# Cardiovascular Outcomes with α1 Adrenergic Receptor Antagonists vs 5α-Reductase Inhibitors

**DOI:** 10.64898/2026.02.23.26346940

**Authors:** LG Kirkland, M Goyal, DJ Kubinski, S Zhuo, Q Li, BC Jensen

## Abstract

**Background:** α-Blockers (ABs) are the most commonly prescribed medications for benign prostatic hyperplasia (BPH), a highly prevalent condition. Although the ALLHAT raised concerns about cardiovascular (CV) safety of nonselective ABs for the treatment of hypertension, the comparative CV risk profile of selective α1A-adrenergic receptor (α1A-AR) antagonists for BPH remains unclear.

**Methods:** We conducted a retrospective cohort study using the TriNetX federated research network (158 million patients across 113 healthcare organizations). Males aged 55 to 90 years with BPH who initiated ABs or 5-ARIs between October 1, 2015, and database lock were included. Three propensity score–matched analyses were conducted: (1) selective α1A-AR antagonists versus 5-ARIs (n=48,096 per group); (2) nonselective ABs versus 5-ARIs (n=33,232 per group); and (3) α1A-selective versus nonselective ABs (n=54,872 per group). Exposures were new use of selective α1A-AR antagonists, nonselective ABs, or 5-ARIs with evidence of adherence. Main outcomes and measures were heart failure (HF) hospitalization, acute MI, stroke, any hospitalization, major adverse CV events (MACE), and composite MACE plus HF at 1, 3, and 5 years.

**Results:** In the α1A-selective AB versus 5-ARI analysis, α1A-selective ABs were associated with increased risk at 1 year of HF (hazard ratio [HR], 1.48 [95% CI, 1.39-1.57]), MI (HR, 1.41 [95% CI, 1.28-1.54]), and stroke (HR, 1.36 [95% CI, 1.22-1.50]). Similar patterns were observed for nonselective ABs versus 5-ARIs: HF (HR, 1.46), MI (HR, 1.29), stroke (HR, 1.32). At 5 years, CV risks remained elevated: HF (HR, 1.50 for selective ABs; HR, 1.52 for nonselective ABs), MI (HR, 1.41; HR, 1.30), and stroke (HR, 1.37; HR, 1.29). Head-to-head comparison of selective versus nonselective ABs showed similar CV outcomes (HF HR, 1.10 at 1 year).

**Conclusions:** Both α1A-selective and nonselective ABs were associated with increased CV event risk compared with 5-ARIs that persisted through 5 years of follow-up. These findings, which were robust across sensitivity analyses and specific to clinically important CV endpoints, may inform shared decision-making for BPH pharmacotherapy.

## Introduction

Benign prostatic hyperplasia (BPH) affects over 94 million men worldwide and prevalence increases substantially with age.^1,2^ In the United States, α1-adrenergic receptor blockers (ABs) are the most commonly prescribed medication class for management of lower urinary tract symptoms associated with BPH--over 5 million prescriptions for tamsulosin alone are dispensed annually in the United States.^3,4^ These medications provide symptomatic relief by relaxing smooth muscle in the prostate and bladder neck through antagonism of α1-adrenergic receptors (α1-ARs).^5–7^

There are three molecular subtypes of α1-ARs with distinct tissue distributions and physiological functions: α1A, α1B and α1D. The α1A-AR subtype, which is blocked preferentially by selective agents such as tamsulosin and silodosin, is expressed not only in the lower urinary tract but also in cardiomyocytes and vascular smooth muscle.^8–10^ Preclinical studies have demonstrated that activation of endogenous cardiac α1A-ARs confers cardioprotective effects, promoting physiologic cardiac hypertrophy and protecting against multiple types of cardiac injury.^11–17,18,19^ Taken together these findings suggest that therapeutic pharmacologic blockade of α1A-ARs could have adverse cardiovascular consequences.

Non-selective ABs were developed as antihypertensives and were among the first line agents for this indication as recently as 2000. However, the landmark Antihypertensive and Lipid-Lowering Treatment to Prevent Heart Attack Trial (ALLHAT) raised significant concerns about the cardiovascular safety of ABs.^20–27^ The doxazosin arm was terminated early after this nonselective AB was associated with a 2-fold increased risk of HF compared with chlorthalidone.^20^ As a result, ABs have been confined to third-line status for treatment of hypertension and rarely are prescribed for this indication.

Subsequent investigations have yielded conflicting results. A meta-analysis by Sousa et al found that ABs were associated with increased risk of acute HF (odds ratio, 1.78 [95% CI, 1.46-2.16]).^28^ However, a study by Jackevicius et al using Veterans Affairs data found that ABs were not associated with adverse outcomes and may have been protective in patients with established HF.^29^ There are no published studies directly comparing cardiovascular outcomes related to α1A-selective and non-selective ABs.

In the present study, we hypothesized that there are distinct patterns of cardiovascular risk for both selective and nonselective ABs compared with 5α-reductase inhibitors (5-ARIs), the second most commonly prescribed medication class for LUTS related to BPH.^3,4,30^ To test this hypothesis, we conducted three parallel analyses comparing: (1) α1A-selective antagonists versus 5-ARIs; (2) nonselective ABs versus 5-ARIs; and (3) α1A-selective versus nonselective ABs in a head-to-head comparison.

## Methods

This report follows the Strengthening the Reporting of Observational Studies in Epidemiology (STROBE) reporting guideline.

### Data Source

This study used clinical data from the TriNetX research network, a federated health research network providing access to deidentified electronic health records from 113 healthcare organizations across the United States comprising 158,340,454 patients.^31–34^ The TriNetX platform aggregates longitudinal clinical data including demographics, diagnoses, procedures, medications, and laboratory values from ambulatory and inpatient encounters. Data queries were conducted using the TriNetX Analytics platform, which provides real-time access to patient-level data while maintaining compliance with the Health Insurance Portability and Accountability Act through a federated query system.^31^ Previous validation studies have demonstrated the reliability of TriNetX data for pharmacoepidemiologic research.^35–38^

### Study Design and Population

We conducted an active comparator, new-user cohort study with three parallel analyses (**Figure 1**).^39–41^ This design minimizes confounding by indication and avoids immortal time bias.^42,43^ We included males aged 55 to 90 years with 1 or more diagnosis codes for BPH (International Classification of Diseases, Tenth Revision, Clinical Modification [ICD-10-CM] code N40) who initiated an AB or 5-ARI. We restricted our study period to prescriptions initiated after October 1, 2015, to ensure all outcomes were captured using ICD-10-CM codes.

**Figure 1.**
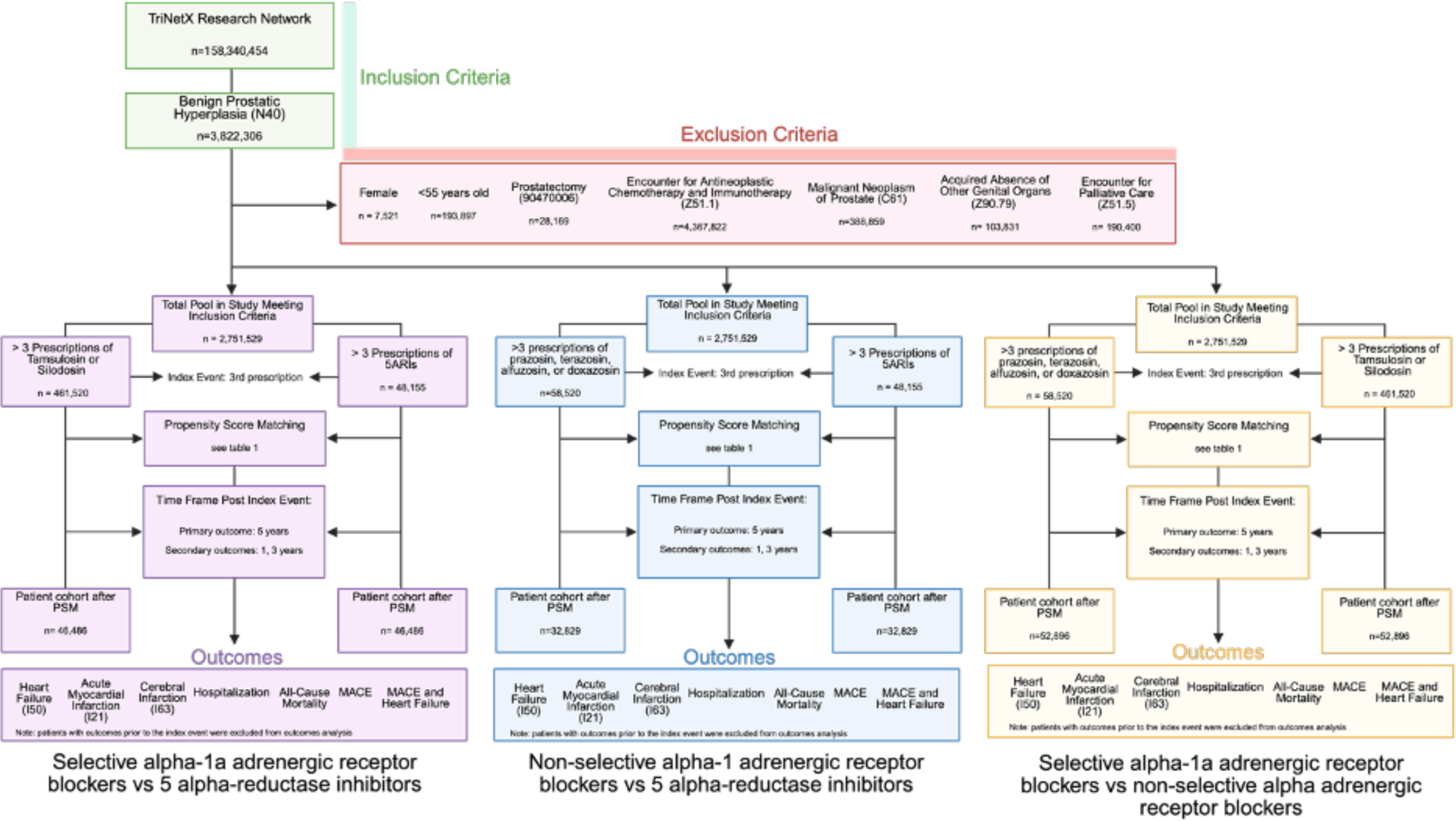
Study Flow Diagram. Schematic representation of patient selection, exclusion criteria, and propensity score matching for three parallel analyses: (1) selective α1A-AR antagonists (tamsulosin, silodosin) versus 5α-reductase inhibitors (5-ARIs); (2) nonselective α-blockers (prazosin, terazosin, alfuzosin, doxazosin) versus 5-ARIs; and (3) α1A-selective versus nonselective α-blockers. The index event was defined as the third prescription fill to ensure medication adherence. PSM indicates propensity score matching.

We excluded patients with prior diagnosis for heart failure (ICD-10-CM codes I50.x), prior myocardial infarction (MI; ICD-10-CM codes I21.x, I22.x), or prior stroke (ICD-10-CM codes I63.x, I64.x) at any time prior to medication initiation. Additional exclusions included: female sex (n = 7,521); age younger than 55 years (n = 193,897); prostatectomy (n = 28,169); encounter for antineoplastic chemotherapy (n = 4,367,822, excluded due to competing mortality risk from cancer); malignant neoplasm of prostate (n = 388,859); acquired absence of other genital organs (n = 103,831); and encounter for palliative care (n = 190,400). After exclusions, 2,751,529 patients met study inclusion criteria.

### Exposure Assessment

We identified study medications using RxNorm codes for prescription orders. Selective α1A-AR antagonists included tamsulosin and silodosin.^44–46^ Nonselective ABs included prazosin, terazosin, alfuzosin, and doxazosin.^47–51^ 5-ARIs include finasteride and dutasteride.^52–57^

To minimize exposure misclassification due to nonadherence and ensure comparability of treatment groups, we required patients to have 3 or more prescriptions for the study drug, with the index event defined as the third prescription fill. This approach ensured that patients demonstrated sustained medication adherence and excluded those who discontinued treatment early. We excluded individuals who filled a prescription for the other drug class or experienced a primary outcome between the first and third prescription.

### Outcomes

Primary outcomes were hospitalization for HF, hospitalization for acute MI, and hospitalization for cerebral infarction (stroke). Secondary outcomes included any hospitalization, major adverse cardiovascular events (MACE; defined as MI, stroke, or death), and composite MACE plus HF. We identified hospitalization outcomes using ICD-10-CM diagnosis codes in any position from inpatient encounters, using algorithms with high specificity (93%-98%) or positive predictive value (>95%).^58–64^ Outcomes were assessed at 1-year, 3-year, and 5-year follow-up intervals.

### Covariates

Covariates were identified through a directed acyclic graph based on clinical knowledge and prior literature. Demographic variables included age (continuous) and race/ethnicity (Black or African American vs other). Race and ethnicity data were included as a proxy for processes of marginalization and structural factors impacting health, not as a biological construct.

Comorbidities included essential hypertension, hyperlipidemia, chronic ischemic heart disease, type 2 diabetes, chronic kidney disease, sleep apnea, cerebrovascular diseases, other anemias, peripheral vascular disease, atherosclerosis, liver disease, other lung disorders, and HIV disease. Baseline medications included β-blockers, aspirin, diuretics, calcium channel blockers, angiotensin-converting enzyme inhibitors, angiotensin II receptor blockers, cardiac vasodilators, hydralazine, and carvedilol. Laboratory values included creatinine, sodium, potassium, hemoglobin, creatinine/urea ratio, and urea.

### Statistical Analysis

We conducted the observational analogue of an intention-to-treat analysis.^65^ Treatment assignment was determined at the index date (third prescription), and patients were retained in their assigned treatment group throughout follow-up, regardless of treatment discontinuation or switching. We aimed to estimate the effect of initiating ABs versus 5-ARIs on incident cardiovascular outcomes. However, because these are nonrandomized observational data and we cannot confirm that all causal criteria were met (i.e. no unmeasured confounding), we report associations throughout.

We used propensity score matching (PSM) to balance measured confounders between treatment groups.^41,66^ The TriNetX platform implements 1:1 greedy nearest-neighbor matching with a caliper width of 0.1 standard deviations of the logit of the propensity score. Propensity scores were estimated using logistic regression modeling the probability of treatment assignment as a function of all measured covariates. We assessed covariate balance using absolute standardized mean differences (SMDs), with SMDs ≤0.1 indicating adequate balance. Baseline demographics prior to PSM are in **eTables 1-3**.

We estimated hazard ratios (HRs) and 95% CIs using Cox proportional hazards regression. HRs are displayed on Kaplan-Meier survival curves at 1-year (365 days), 3-year (1,095 days), and 5-year (1,825 days) timepoints. Although the goal of this study was to estimate effect sizes rather than to test null hypotheses, we report 95% CIs to indicate precision of estimates.

Three prespecified analyses were conducted: Analysis 1 compared selective α1A-AR antagonists (tamsulosin, silodosin) with 5-ARIs; Analysis 2 compared nonselective ABs (prazosin, terazosin, alfuzosin, doxazosin) with 5-ARIs; Analysis 3 compared α1A-selective with nonselective ABs in a head-to-head comparison.

### Sensitivity Analyses

To assess the robustness of our findings to varying definitions of medication adherence, we conducted sensitivity analyses using 1, 3, 7, and 12 prescription thresholds for cohort entry (**eFigure 1**). To evaluate whether observed associations were specific to cardiovascular endpoints or reflected systematic differences in healthcare utilization or ascertainment, we examined two negative control outcomes: sensorineural hearing loss and nonmelanoma skin cancer.^67^ These conditions share similar diagnostic pathways and healthcare contact patterns but lack biological plausibility for association with α1-AR blockade (**eFigure 2**). Additionally, we examined erectile dysfunction and phosphodiesterase-5 inhibitor prescriptions as exploratory secondary outcomes given their clinical relevance to the BPH population (**eFigure 3**).

**Figure 2.**
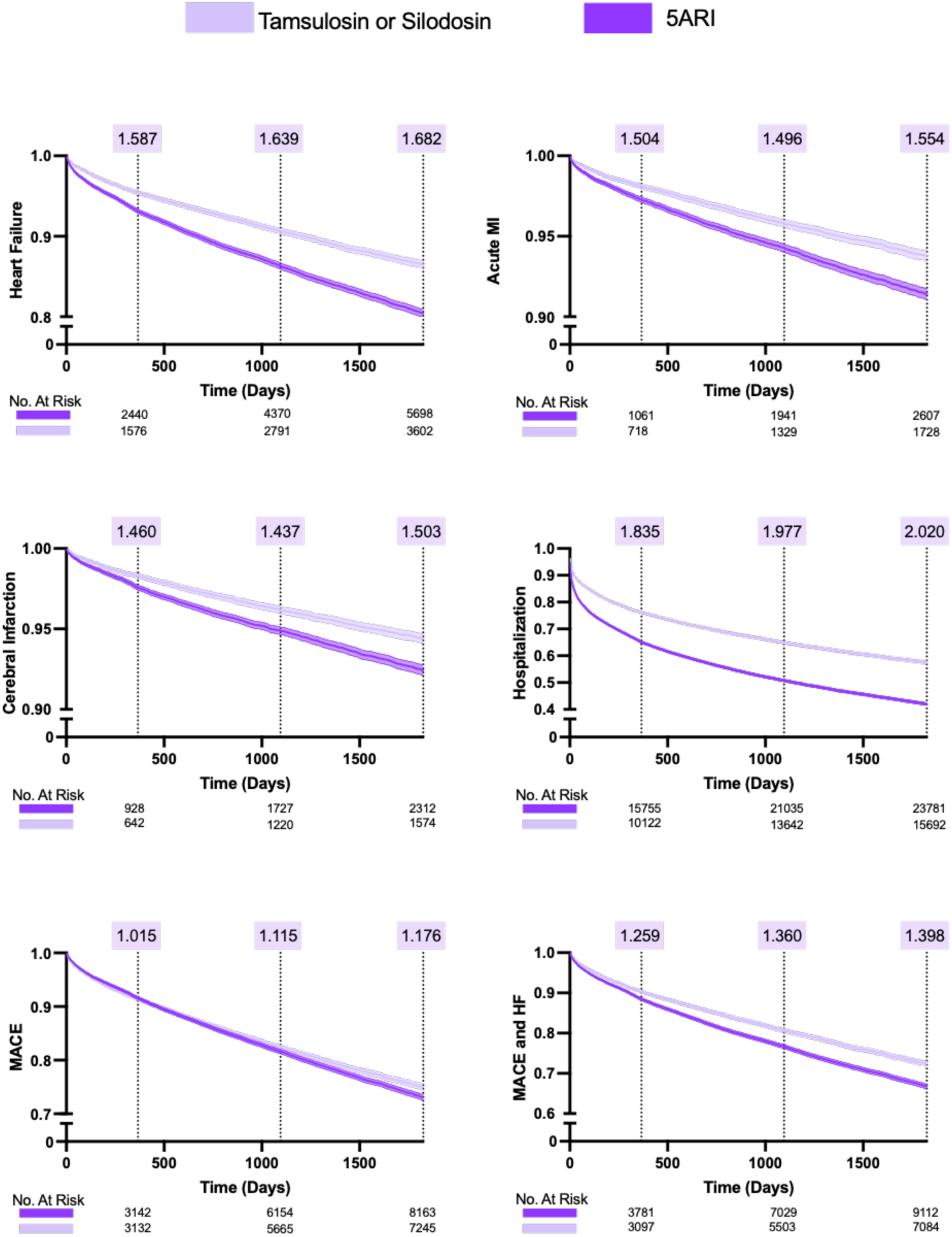
Analysis 1: Selective α-Blockers vs 5-α Reductase Inhibitors. Kaplan-Meier curves for heart failure hospitalization, acute myocardial infarction, cerebral infarction (stroke), all-cause mortality, hospitalization, major adverse cardiovascular events (MACE), and MACE plus heart failure. Hazard ratios displayed at 1-year (365 days), 3-year (1,095 days), and 5-year (1,825 days) timepoints.

**Figure 3.**
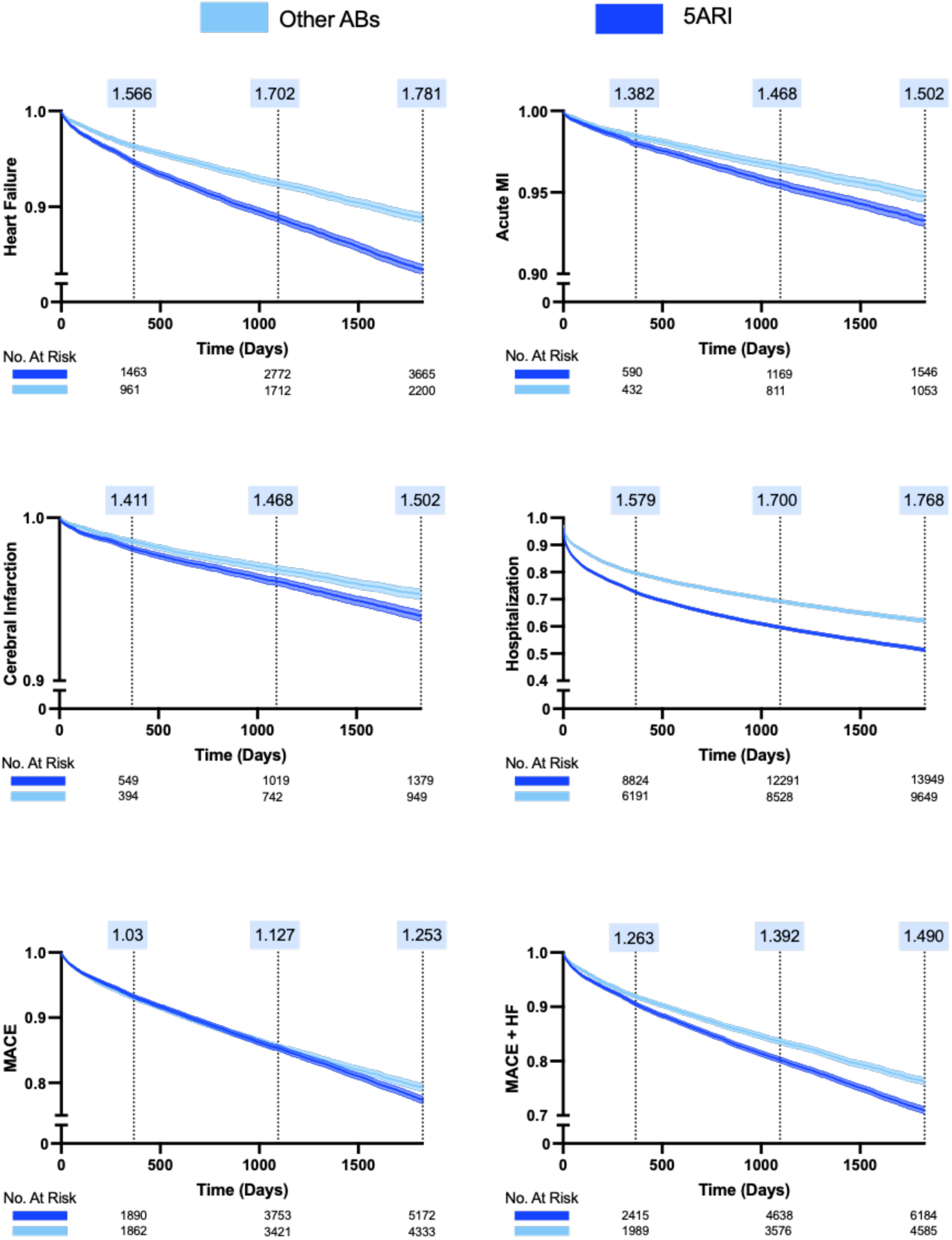
Analysis 2: Nonselective α-Blockers vs 5α- Reductase Inhibitors. Survival curves comparing nonselective α-blockers (prazosin, terazosin, alfuzosin, doxazosin) with 5-α reductase inhibitors. Hazard ratios displayed at 1-year, 3-year, and 5-year timepoints.

## Results

### Study Population

From the TriNetX research network of 158,340,454 patients, we identified 3,822,306 patients with a diagnosis of BPH. After applying exclusion criteria, 2,751,529 patients met initial eligibility requirements. Among these, 486,285 had 3 or more prescriptions for tamsulosin or silodosin, 48,098 had 3 or more prescriptions for 5-ARIs, and 53,872 had 3 or more prescriptions for nonselective ABs (prazosin, terazosin, alfuzosin, or doxazosin).

After propensity score matching, the final cohorts included: Analysis 1 (α1A-selective ABs vs 5-ARIs), 48,096 patients per group; Analysis 2 (nonselective ABs vs 5-ARIs), 33,232 patients per group; and Analysis 3 (α1A-selective vs nonselective ABs), 54,872 patients per group (**Figure 1**).

### Baseline Characteristics

After propensity score matching, baseline characteristics were well balanced between treatment groups in all three analyses (**Tables 1-3**). In Analysis 1 (α1A-selective ABs vs 5-ARIs), mean age was 72.99 years (SD, 9.90) in the selective AB group and 73.68 years (SD, 9.99) in the 5-ARI group. The cohorts were predominantly non-Black (approximately 94%), with balanced distributions of cardiovascular risk factors including essential hypertension (62.80% vs 62.41%), hyperlipidemia (50.59% vs 49.26%), chronic ischemic heart disease (30.09% vs 31.00%), type 2 diabetes (24.95% vs 25.86%), and chronic kidney disease (17.35% vs 18.00%). Baseline β-blocker use was similar (48.36% vs 47.19%). All SMDs were ≤0.10 after matching, indicating excellent covariate balance.

**Table 1.** Baseline Characteristics of the Selective α1A-AR Antagonists and 5α-Reductase Inhibitors Groups After Propensity Score Matching.

|  |  | Cohort Statistics After PSM |  |  |  |  |  |  |  |
| --- | --- | --- | --- | --- | --- | --- | --- | --- | --- |
| Category | Characteristic | Selective $\alpha$ -Blockers | | | 5 - $\alpha$ - Reductase Inhibitors | | | p-value | SMD |
|  |  | Count(%) | Mean | SD | Count(%) | Mean | SD |  |  |
| Demographics | Age at Index | 48096 (100%) | 72.992 | 9.903 | 48096 (100%) | 73.679 | 9.999 | <0.0001 | 0.069 |
|  | Black or African American | 2700 (5.61%) | – | – | 3143 (6.54%) | – | – | <0.0001 | 0.039 |
| Diseases | Essential hypertension | 30202 (62.80%) | – | – | 30015 (62.41%) | – | – | <0.0001 | 0.008 |
|  | Hyperlipidemia | 24331 (50.59%) | – | – | 23690 (49.26%) | – | – | <0.0001 | 0.027 |
|  | Chronic ischemic heart disease | 14470 (30.09%) | – | – | 14910 (31.00%) | – | – | 0.002 | 0.020 |
|  | Type 2 diabetes | 11998 (24.95%) | – | – | 12439 (25.86%) | – | – | 0.001 | 0.021 |
|  | Chronic kidney disease | 8345 (17.35%) | – | – | 8657 (18.00%) | – | – | 0.008 | 0.017 |
|  | Sleep apnea | 7199 (14.97%) | – | – | 7146 (14.86%) | – | – | 0.631 | 0.003 |
|  | Other anemias | 7059 (14.68%) | – | – | 7332 (15.25%) | – | – | 0.014 | 0.016 |
|  | Cerebrovascular diseases | 7049 (14.66%) | – | – | 7293 (15.16%) | – | – | 0.027 | 0.014 |
|  | Atherosclerosis | 3130 (6.51%) | – | – | 3240 (6.74%) | – | – | 0.154 | 0.009 |
|  | Peripheral vascular disease | 3105 (6.46%) | – | – | 3260 (6.78%) | – | – | 0.044 | 0.013 |
|  | Liver disease | 2661 (5.53%) | – | – | 2780 (5.78%) | – | – | 0.097 | 0.011 |
|  | Other lung disorders | 1697 (3.53%) | – | – | 1761 (3.66%) | – | – | 0.268 | 0.007 |
|  | HIV disease | 225 (0.47%) | – | – | 268 (0.56%) | – | – | 0.052 | 0.013 |
| Medications | Beta blockers | 23259 (48.36%) | – | – | 22697 (47.19%) | – | – | <0.0001 | 0.023 |
|  | Aspirin | 19096 (39.70%) | – | – | 18530 (38.53%) | – | – | <0.0001 | 0.024 |
|  | Diuretics | 18093 (37.62%) | – | – | 17585 (36.56%) | – | – | <0.0001 | 0.022 |
|  | Calcium channel blockers | 15509 (32.25%) | – | – | 14832 (30.84%) | – | – | <0.0001 | 0.030 |
|  | ACE inhibitors | 14919 (31.02%) | – | – | 13951 (29.01%) | – | – | <0.0001 | 0.044 |
|  | Angiotensin II inhibitors | 11332 (23.56%) | – | – | 10576 (21.99%) | – | – | <0.0001 | 0.037 |
|  | Vasodilators (cardiac) | 7247 (15.07%) | – | – | 7353 (15.29%) | – | – | 0.341 | 0.006 |
|  | Hydralazine | 5829 (12.12%) | – | – | 5944 (12.36%) | – | – | 0.258 | 0.007 |
|  | Carvedilol | 3957 (8.23%) | – | – | 4156 (8.64%) | – | – | 0.021 | 0.015 |
| Lab Values | Creatinine | 38219 (79.46%) | 1.249 | 1.973 | 37649 (78.28%) | 1.236 | 2.226 | 0.398 | 0.006 |
|  | Sodium | 37764 (78.52%) | 138.579 | 3.717 | 37266 (77.48%) | 138.819 | 3.450 | <0.0001 | 0.067 |
|  | Potassium | 37552 (78.08%) | 4.215 | 0.491 | 37019 (76.97%) | 4.210 | 0.472 | 0.197 | 0.009 |
|  | Hemoglobin | 35859 (74.56%) | 13.192 | 2.325 | 35384 (73.57%) | 13.093 | 2.296 | <0.0001 | 0.043 |
|  | Creatinine/Urea ratio | 1169 (2.43%) | 17.762 | 6.138 | 1248 (2.60%) | 18.866 | 6.760 | <0.0001 | 0.171 |
|  | Urea | 219 (0.46%) | 25.602 | 19.373 | 208 (0.43%) | 28.441 | 17.707 | 0.1152 | 0.153 |

**Table 2.**
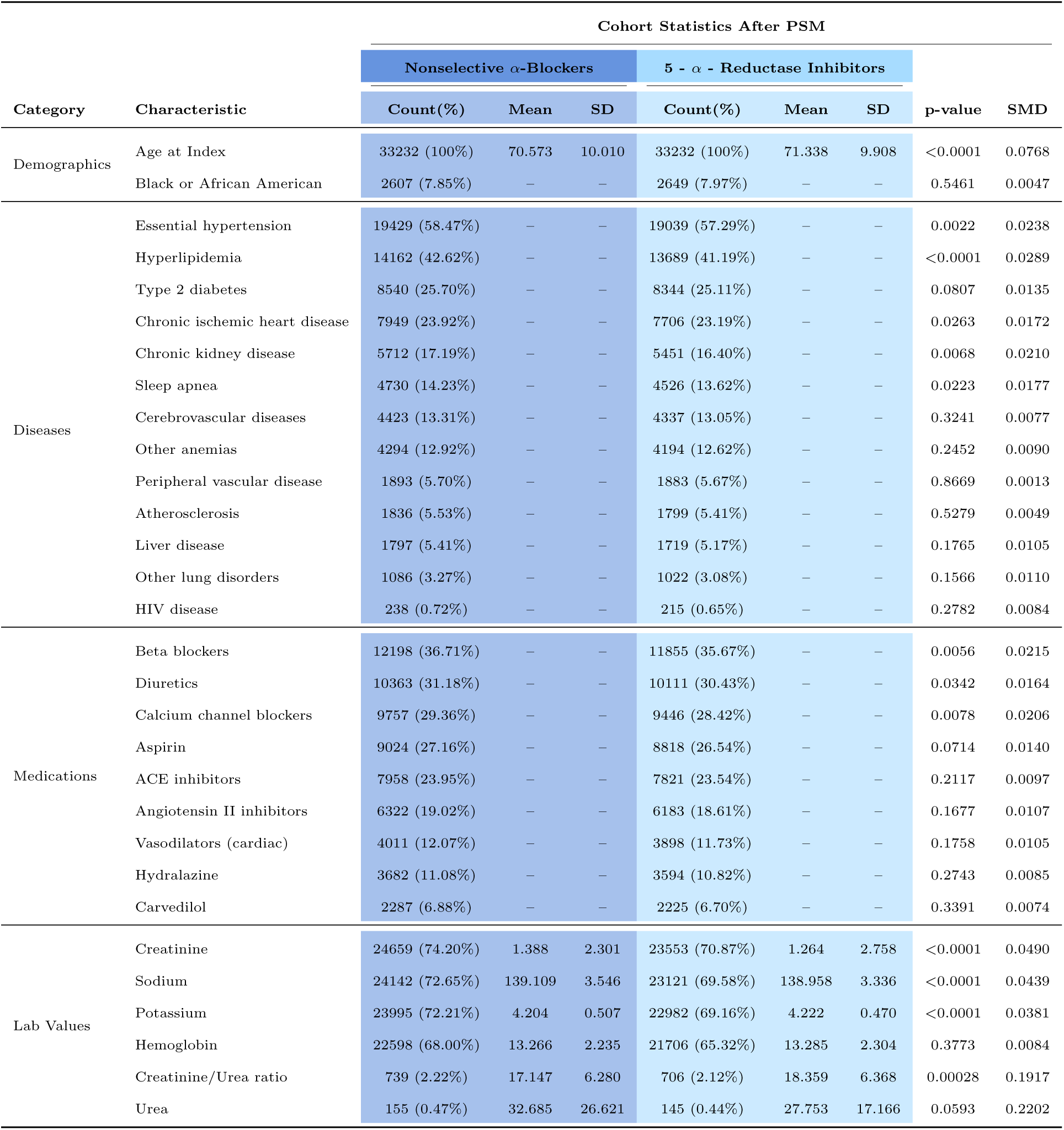
Baseline Characteristics of the Nonselective α-Blockers and 5α-Reductase Inhibitors Groups After Propensity Score Matching.

**Table 3.** Baseline Characteristics of the α1A-Selective and Nonselective α-Blockers Groups After Propensity Score Matching.

| Category | Characteristic | Cohort Statistics After PSM |  |  |  |  |  |  |  |
| --- | --- | --- | --- | --- | --- | --- | --- | --- | --- |
| | | α1A-Selective | | | Nonselective α-Blockers | | | p-value | SMD |
|  |  | Count(%) | Mean | SD | Count(%) | Mean | SD |  |  |
| Demographics | Age at Index | 54872 (100%) | 69.083 | 10.089 | 54872 (100%) | 68.847 | 10.233 | 0.0001 | 0.023 |
|  | Black or African American | 5276 (9.62%) | – | – | 5653 (10.30%) | – | – | 0.0001 | 0.023 |
| Diseases | Essential hypertension | 27888 (50.82%) | – | – | 28368 (51.70%) | – | – | 0.004 | 0.018 |
|  | Hyperlipidemia | 17703 (32.26%) | – | – | 17977 (32.76%) | – | – | <0.0001 | 0.011 |
|  | Type 2 diabetes | 12282 (22.38%) | – | – | 12867 (23.45%) | – | – | <0.0001 | 0.025 |
|  | Chronic ischemic heart disease | 9807 (17.87%) | – | – | 10131 (18.46%) | – | – | 0.011 | 0.015 |
|  | Chronic kidney disease | 7403 (13.49%) | – | – | 8199 (14.94%) | – | – | <0.0001 | 0.042 |
|  | Sleep apnea | 5973 (10.89%) | – | – | 6334 (11.54%) | – | – | <0.0001 | 0.021 |
|  | Cerebrovascular diseases | 5604 (10.21%) | – | – | 5922 (10.79%) | – | – | 0.002 | 0.019 |
|  | Other anemias | 4916 (8.96%) | – | – | 5440 (9.91%) | – | – | <0.0001 | 0.033 |
|  | Peripheral vascular disease | 2233 (4.07%) | – | – | 2470 (4.50%) | – | – | 0.0004 | 0.021 |
|  | Liver disease | 2129 (3.88%) | – | – | 2301 (4.19%) | – | – | 0.0083 | 0.016 |
|  | Atherosclerosis | 2027 (3.69%) | – | – | 2261 (4.12%) | – | – | 0.0003 | 0.022 |
|  | Other lung disorders | 1225 (2.23%) | – | – | 1353 (2.47%) | – | – | 0.0107 | 0.015 |
|  | HIV disease | 331 (0.60%) | – | – | 390 (0.71%) | – | – | 0.0275 | 0.013 |
| Medications | Beta blockers | 13217 (24.09%) | – | – | 13975 (25.47%) | – | – | <0.0001 | 0.032 |
|  | Diuretics | 11693 (21.31%) | – | – | 12458 (22.70%) | – | – | <0.0001 | 0.034 |
|  | Calcium channel blockers | 11307 (20.61%) | – | – | 12278 (22.38%) | – | – | <0.0001 | 0.043 |
|  | Aspirin | 9665 (17.61%) | – | – | 9970 (18.17%) | – | – | 0.0163 | 0.015 |
|  | ACE inhibitors | 8689 (15.84%) | – | – | 9142 (16.66%) | – | – | 0.0002 | 0.022 |
|  | Angiotensin II inhibitors | 6832 (12.45%) | – | – | 7429 (13.54%) | – | – | <0.0001 | 0.032 |
|  | Vasodilators (cardiac) | 4377 (7.98%) | – | – | 4797 (8.74%) | – | – | <0.0001 | 0.028 |
|  | Hydralazine | 3904 (7.12%) | – | – | 4464 (8.14%) | – | – | <0.0001 | 0.038 |
|  | Carvedilol | 2249 (4.10%) | – | – | 2676 (4.88%) | – | – | <0.0001 | 0.038 |
| Lab Values | Creatinine | 31749 (57.86%) | 1.281 | 1.906 | 31836 (58.02%) | 1.432 | 2.787 | <0.0001 | 0.063 |
|  | Sodium | 30546 (55.67%) | 138.591 | 3.719 | 30564 (55.70%) | 139.066 | 3.532 | <0.0001 | 0.131 |
|  | Potassium | 30517 (55.62%) | 4.204 | 0.498 | 30541 (55.66%) | 4.200 | 0.509 | 0.302 | 0.008 |
|  | Hemoglobin | 28777 (52.44%) | 13.372 | 2.292 | 28845 (52.57%) | 13.283 | 2.235 | <0.0001 | 0.039 |
|  | Creatinine/Urea ratio | 822 (1.50%) | 17.269 | 6.155 | 900 (1.64%) | 16.736 | 6.151 | 0.0729 | 0.087 |
|  | Urea | 189 (0.34%) | 26.049 | 19.572 | 187 (0.34%) | 32.399 | 25.606 | 0.0072 | 0.279 |

In Analysis 2 (nonselective ABs vs 5-ARIs), mean age was 70.57 years (SD, 10.01) versus 71.34 years (SD, 9.91). Cardiovascular comorbidities were similarly balanced. In Analysis 3 (selective vs nonselective ABs), mean age was 68.85 years (SD, 10.09) in both groups after matching.

### Analysis 1: Selective α1A-AR Antagonists vs 5α-Reductase Inhibitors

At 1 year, α1A-selective ABs (tamsulosin, silodosin) compared with 5-ARIs were associated with increased risk of HF hospitalization (HR, 1.48 [95% CI, 1.39-1.57]), acute MI (HR, 1.41 [95% CI, 1.28-1.54]), and cerebral infarction (HR, 1.36 [95% CI, 1.22-1.50]) (**Figure 2**, **Table 4**). Risk of hospitalization for any cause was substantially higher with α1A-selective ABs (HR, 1.58 [95% CI, 1.54-1.62]).

**Table 4.** Selective α1A-AR antagonists vs. 5α-Reductase Inhibitors Primary and Secondary Outcomes.

| Outcome | RD (95% CI) | z | p | RR (95% CI) | OR (95% CI) | HR (95% CI) |
| --- | --- | --- | --- | --- | --- | --- |
| Heart Failure | 2.19% ± 0.30% | 14.057 | <0.0001 | 1.55 ± 0.10 | 1.59 ± 0.10 | 1.48 ± 0.09 |
| Acute MI | 0.80% ± 0.19% | 8.414 | <0.0001 | 1.49 ± 0.14 | 1.50 ± 0.15 | 1.41 ± 0.13 |
| Cerebral Infarction | 0.65% ± 0.17% | 7.346 | <0.0001 | 1.45 ± 0.15 | 1.46 ± 0.15 | 1.36 ± 0.14 |
| Hospitalization | 11.93% ± 0.56% | 41.097 | <0.0001 | 1.56 ± 0.03 | 1.84 ± 0.05 | 1.58 ± 0.04 |
| MACE | 0.11% ± 0.36% | 0.566 | 0.5713 | 1.01 ± 0.05 | 1.02 ± 0.05 | 0.96 ± 0.05 |
| MACE + HF | 2.00% ± 0.43% | 9.066 | <0.0001 | 1.23 ± 0.06 | 1.26 ± 0.06 | 1.18 ± 0.06 |

| 3 Year Outcome |  |  |  |  |  |  |
| --- | --- | --- | --- | --- | --- | --- |
| Outcome | RD (95% CI) | z | p | RR (95% CI) | OR (95% CI) | HR (95% CI) |
| Heart Failure | 3.99% ± 0.40% | 19.650 | <0.0001 | 1.57 ± 0.07 | 1.64 ± 0.08 | 1.48 ± 0.07 |
| Acute MI | 1.43% ± 0.25% | 11.178 | <0.0001 | 1.47 ± 0.10 | 1.50 ± 0.11 | 1.36 ± 0.10 |
| Cerebral Infarction | 1.16% ± 0.24% | 9.586 | <0.0001 | 1.42 ± 0.10 | 1.44 ± 0.11 | 1.31 ± 0.10 |
| Hospitalization | 15.66% ± 0.61% | 49.905 | <0.0001 | 1.54 ± 0.03 | 1.98 ± 0.05 | 1.58 ± 0.03 |
| MACE | 1.35% ± 0.48% | 5.482 | <0.0001 | 1.10 ± 0.04 | 1.12 ± 0.04 | 1.02 ± 0.04 |
| MACE + HF | 4.43% ± 0.55% | 15.563 | <0.0001 | 1.29 ± 0.04 | 1.36 ± 0.05 | 1.22 ± 0.04 |

**Table 4. Selective α1A-AR antagonists vs. 5α-Reductase Inhibitors Primary and Secondary Outcomes.**
| 5 Year Outcome |  |  |  |  |  |  |
| --- | --- | --- | --- | --- | --- | --- |
| Outcome | RD (95% CI) | z | p | RR (95% CI) | OR (95% CI) | HR (95% CI) |
| Heart Failure | 5.23% ± 0.44% | 23.238 | <0.0001 | 1.59 ± 0.06 | 1.68 ± 0.07 | 1.50 ± 0.06 |
| Acute MI | 2.02% ± 0.28% | 13.976 | <0.0001 | 1.52 ± 0.09 | 1.55 ± 0.10 | 1.41 ± 0.09 |
| Cerebral Infarction | 1.67% ± 0.27% | 12.283 | <0.0001 | 1.48 ± 0.09 | 1.50 ± 0.10 | 1.37 ± 0.09 |
| Hospitalization | 16.82% ± 0.61% | 53.021 | <0.0001 | 1.52 ± 0.02 | 2.02 ± 0.05 | 1.57 ± 0.03 |
| MACE | 2.44% ± 0.53% | 9.052 | <0.0001 | 1.14 ± 0.03 | 1.18 ± 0.04 | 1.06 ± 0.03 |
| MACE + HF | 5.79% ± 0.61% | 18.699 | <0.0001 | 1.30 ± 0.04 | 1.40 ± 0.05 | 1.24 ± 0.04 |

At 3 years, the pattern of increased cardiovascular events persisted for HF (HR, 1.48 [95% CI, 1.41-1.55]), MI (HR, 1.36 [95% CI, 1.26-1.46]), stroke (HR, 1.31 [95% CI, 1.21-1.41]), and hospitalization (HR, 1.58 [95% CI, 1.55-1.61]). MACE plus HF increased (HR, 1.22 [95% CI, 1.18-1.26]).

At 5 years, cardiovascular event risks associated with selective α1A antagonists vs. 5-ARIs remained elevated: HF (HR, 1.50 [95% CI, 1.44-1.56]), MI (HR, 1.41 [95% CI, 1.32-1.50]), stroke (HR, 1.37 [95% CI, 1.28-1.46]), and hospitalization (HR, 1.57 [95% CI, 1.54-1.60]). MACE was modestly elevated (HR, 1.06 [95% CI, 1.03-1.09]), and MACE plus HF showed sustained elevation (HR, 1.24 [95% CI, 1.20-1.28]).

### Analysis 2: Nonselective α-Blockers vs 5α-Reductase Inhibitors

Nonselective ABs (prazosin, terazosin, alfuzosin, doxazosin) demonstrated similar cardiovascular risk patterns when compared with 5-ARIs as α1A-selective ABs (**Figure 3**, **Table 5**). At 1 year, nonselective ABs were associated with increased risk of HF (HR, 1.46 [95% CI, 1.34-1.58]), MI (HR, 1.29 [95% CI, 1.13-1.45]), stroke (HR, 1.32 [95% CI, 1.15-1.49]), and hospitalization (HR, 1.42 [95% CI, 1.37-1.47]).

**Table 5.** Nonselective α-Blockers vs 5α-Reductase Inhibitors Primary and Secondary Outcomes.

| 1 Year Outcome |  |  |  |  |  |  |
| --- | --- | --- | --- | --- | --- | --- |
| Outcome | RD (95% CI) | z | p | RR (95% CI) | OR (95% CI) | HR (95% CI) |
| Heart Failure | 1.75% ± 0.32% | 10.67 | <0.0001 | 1.54 ± 0.12 | 1.57 ± 0.13 | 1.46 ± 0.12 |
| Acute MI | 0.51% ± 0.20% | 5.09 | <0.0001 | 1.37 ± 0.17 | 1.38 ± 0.17 | 1.29 ± 0.16 |
| Cerebral Infarction | 0.51% ± 0.19% | 5.20 | <0.0001 | 1.40 ± 0.18 | 1.41 ± 0.18 | 1.32 ± 0.17 |
| Hospitalization | 7.92% ± 0.63% | 24.42 | <0.0001 | 1.43 ± 0.04 | 1.58 ± 0.06 | 1.42 ± 0.05 |
| MACE | 0.17% ± 0.39% | 0.87 | 0.3833 | 1.03 ± 0.06 | 1.03 ± 0.07 | 0.97 ± 0.06 |
| MACE + HF | 1.74% ± 0.46% | 7.42 | <0.0001 | 1.24 ± 0.07 | 1.26 ± 0.08 | 1.18 ± 0.07 |

| 3 Year Outcome |  |  |  |  |  |  |
| --- | --- | --- | --- | --- | --- | --- |
| Outcome | RD (95% CI) | z | p | RR (95% CI) | OR (95% CI) | HR (95% CI) |
| Heart Failure | 3.69% ± 0.43% | 16.83 | <0.0001 | 1.64 ± 0.10 | 1.70 ± 0.11 | 1.50 ± 0.09 |
| Acute MI | 1.16% ± 0.27% | 8.32 | <0.0001 | 1.45 ± 0.13 | 1.47 ± 0.13 | 1.30 ± 0.12 |
| Cerebral Infarction | 0.90% ± 0.26% | 6.85 | <0.0001 | 1.38 ± 0.13 | 1.40 ± 0.14 | 1.25 ± 0.12 |
| Hospitalization | 11.32% ± 0.70% | 31.47 | <0.0001 | 1.44 ± 0.03 | 1.70 ± 0.06 | 1.41 ± 0.04 |
| MACE | 1.27% ± 0.52% | 4.75 | <0.0001 | 1.11 ± 0.05 | 1.13 ± 0.06 | 1.00 ± 0.05 |
| MACE + HF | 4.24% ± 0.60% | 13.75 | <0.0001 | 1.32 ± 0.06 | 1.39 ± 0.07 | 1.22 ± 0.05 |

**Table 5. Nonselective α-Blockers vs 5α-Reductase Inhibitors Primary and Secondary Outcomes.**
| 5 Year Outcome |  |  |  |  |  |  |
| --- | --- | --- | --- | --- | --- | --- |
| Outcome | RD (95% CI) | z | p | RR (95% CI) | OR (95% CI) | HR (95% CI) |
| Heart Failure | 5.09% ± 0.48% | 20.58 | <0.0001 | 1.68 ± 0.09 | 1.78 ± 0.10 | 1.52 ± 0.08 |
| Acute MI | 1.59% ± 0.31% | 10.04 | <0.0001 | 1.48 ± 0.11 | 1.50 ± 0.12 | 1.30 ± 0.10 |
| Cerebral Infarction | 1.40% ± 0.30% | 9.26 | <0.0001 | 1.46 ± 0.12 | 1.48 ± 0.13 | 1.29 ± 0.11 |
| Hospitalization | 12.94% ± 0.72% | 34.86 | <0.0001 | 1.45 ± 0.03 | 1.77 ± 0.06 | 1.41 ± 0.04 |
| MACE | 3.02% ± 0.59% | 10.06 | <0.0001 | 1.21 ± 0.04 | 1.25 ± 0.06 | 1.07 ± 0.04 |
| MACE + HF | 6.33% ± 0.67% | 18.42 | <0.0001 | 1.38 ± 0.05 | 1.49 ± 0.06 | 1.25 ± 0.05 |

At 3 years, cardiovascular event risks remained elevated: HF (HR, 1.50 [95% CI, 1.41-1.59]), MI (HR, 1.30 [95% CI, 1.18-1.42]), stroke (HR, 1.25 [95% CI, 1.13-1.37]), and hospitalization (HR, 1.41 [95% CI, 1.37-1.45]) were higher in the non-selective AB than the 5-ARI group.

At 5 years, cardiovascular events persisted: HF (HR, 1.52 [95% CI, 1.44-1.60]), MI (HR, 1.30 [95% CI, 1.20-1.40]), stroke (HR, 1.29 [95% CI, 1.18-1.40]), and hospitalization (HR, 1.41 [95% CI, 1.37-1.45]) were higher in the non-selective AB than the 5-ARI group.

### Analysis 3: α1A-Selective vs Nonselective α-Blockers

Direct comparison of selective versus nonselective ABs revealed generally similar cardiovascular outcomes (**Figure 4**, **Table 6**). At 1 year, there were modestly higher cardiovascular risks in the α1A-selective AB group for HF (HR, 1.10 [95% CI, 1.03-1.16]), MI (HR, 1.16 [95% CI, 1.06-1.27]), stroke (HR, 1.28 [95% CI, 1.16-1.40]), and hospitalization (HR, 1.26 [95% CI, 1.23-1.29]).

**Figure 4.**
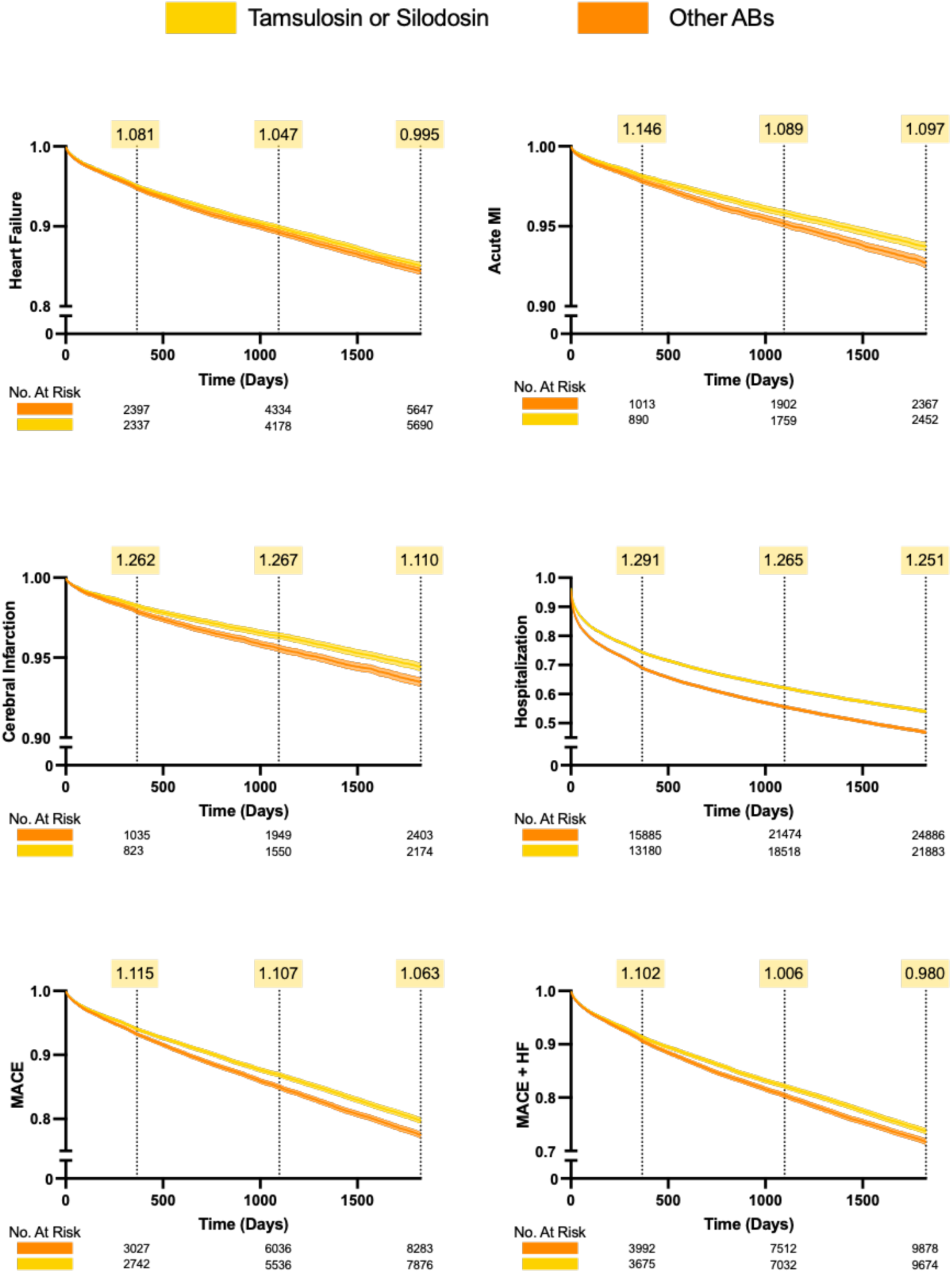
Analysis 3: α1A-Selective vs Nonselective α-Blockers. Head-to-head comparison of selective (tamsulosin, silodosin) versus nonselective (prazosin, terazosin, alfuzosin, doxazosin) α-blockers. Hazard ratios displayed at 1-year, 3-year, and 5-year timepoints.

**Table 6.** Selective α1A-AR Antagonists and Nonselective α-Blockers Primary and Secondary Outcomes.

| 1 Year Outcome |  |  |  |  |  |  |
| --- | --- | --- | --- | --- | --- | --- |
| Outcome | RD (95% CI) | z | p | RR (95% CI) | OR (95% CI) | HR (95% CI) |
| Heart Failure | 0.36% ± 0.27% | 2.593 | 0.0095 | 1.077 ± 0.061 | 1.081 ± 0.064 | 1.095 ± 0.064 |
| Acute MI | 0.25% ± 0.17% | 2.941 | 0.0033 | 1.143 ± 0.102 | 1.146 ± 0.105 | 1.163 ± 0.105 |
| Cerebral Infarction | 0.41% ± 0.16% | 4.955 | <0.0001 | 1.257 ± 0.114 | 1.262 ± 0.117 | 1.279 ± 0.117 |
| Hospitalization | 5.02% ± 0.53% | 18.567 | <0.0001 | 1.205 ± 0.024 | 1.291 ± 0.035 | 1.257 ± 0.029 |
| MACE | 0.60% ± 0.30% | 3.992 | <0.0001 | 1.107 ± 0.056 | 1.115 ± 0.060 | 1.128 ± 0.058 |
| MACE + HF | 0.75% ± 0.36% | 4.052 | <0.0001 | 1.093 ± 0.047 | 1.102 ± 0.052 | 1.112 ± 0.050 |

| 3 Year Outcome |  |  |  |  |  |  |
| --- | --- | --- | --- | --- | --- | --- |
| Outcome | RD (95% CI) | z | p | RR (95% CI) | OR (95% CI) | HR (95% CI) |
| Heart Failure | 0.37% ± 0.36% | 2.026 | 0.0427 | 1.043 ± 0.042 | 1.047 ± 0.046 | 1.080 ± 0.046 |
| Acute MI | 0.29% ± 0.23% | 2.539 | 0.0111 | 1.086 ± 0.069 | 1.089 ± 0.072 | 1.130 ± 0.074 |
| Cerebral Infarction | 0.78% ± 0.22% | 6.852 | <0.0001 | 1.257 ± 0.083 | 1.267 ± 0.086 | 1.312 ± 0.088 |
| Hospitalization | 5.49% ± 0.58% | 18.64 | <0.0001 | 1.160 ± 0.018 | 1.265 ± 0.032 | 1.236 ± 0.024 |
| MACE | 1.06% ± 0.40% | 5.131 | <0.0001 | 1.094 ± 0.038 | 1.107 ± 0.043 | 1.144 ± 0.042 |
| MACE + HF | 1.16% ± 0.48% | 4.73 | <0.0001 | 1.075 ± 0.032 | 1.089 ± 0.039 | 1.120 ± 0.037 |

**Table 6. Selective α1A-AR Antagonists and Nonselective α-Blockers Primary and Secondary Outcomes.**
| 5 Year Outcome |  |  |  |  |  |  |
| --- | --- | --- | --- | --- | --- | --- |
| Outcome | RD (95% CI) | z | p | RR (95% CI) | OR (95% CI) | HR (95% CI) |
| Heart Failure | -0.05% ± 0.40% | -0.232 | 0.8164 | 0.996 ± 0.035 | 0.995 ± 0.039 | 1.056 ± 0.039 |
| Acute MI | 0.43% ± 0.26% | 3.242 | 0.0012 | 1.092 ± 0.058 | 1.097 ± 0.062 | 1.171 ± 0.064 |
| Cerebral Infarction | 0.44% ± 0.25% | 3.445 | 0.0006 | 1.105 ± 0.062 | 1.110 ± 0.066 | 1.185 ± 0.069 |
| Hospitalization | 5.47% ± 0.58% | 18.331 | <0.0001 | 1.137 ± 0.016 | 1.251 ± 0.030 | 1.236 ± 0.023 |
| MACE | 0.83% ± 0.46% | 3.562 | 0.0004 | 1.053 ± 0.030 | 1.063 ± 0.036 | 1.136 ± 0.035 |
| MACE + HF | 0.52% ± 0.53% | 1.937 | 0.0528 | 1.025 ± 0.026 | 1.032 ± 0.033 | 1.095 ± 0.031 |

Collectively, these findings suggest that cardiovascular outcome differences observed in Analyses 1 and 2 are attributable primarily to differences between all ABs and 5-ARIs, rather than to differences between α1A-selective and nonselective AB subclasses.

### Sensitivity and Secondary Analyses

Sensitivity analyses varying the prescription threshold for cohort entry (1, 3, 7, and 12 prescriptions) demonstrated consistent patterns across all thresholds, with cardiovascular event rates increasing as the prescription requirement increased (**eFigure 1**). This pattern is consistent with healthy user bias at lower thresholds and survivor bias at higher thresholds, with our primary analysis (3 prescriptions) representing a reasonable balance.

Negative control outcome analyses revealed no significant differences in sensorineural hearing loss or skin cancer between treatment groups across all three comparisons (**eFigure 2**).^67^ The absence of spurious associations with these negative control outcomes supports the specificity of our cardiovascular findings and argues against systematic ascertainment bias or unmeasured confounding affecting all outcomes equally.

Exploratory analyses of erectile dysfunction showed selective ABs were associated with modestly increased risk compared with 5-ARIs (HR, 1.32 at 1 year) and compared with nonselective ABs (HR, 0.98 at 1 year), while phosphodiesterase-5 inhibitor prescriptions showed similar patterns (**eFigure 3**). These findings may reflect differences in patient populations or healthcare engagement patterns between treatment groups.

## Discussion

In this large retrospective cohort study of propensity score–matched patients with BPH initiating pharmacotherapy, we found that both selective α1A-AR antagonists (tamsulosin, silodosin) and nonselective ABs were associated with increased risk of HF, MI, stroke, and any hospitalization compared with 5-ARIs. These elevated cardiovascular risks were evident at 1 year and persisted through 5 years of follow-up. Sensitivity analyses using varying prescription thresholds and negative control outcomes supported the robustness and specificity of these findings.

Our findings demonstrate that antagonism of the α1A-AR subtype is sufficient to increase cardiovascular risk and that activity at the other α1-AR subtypes likely does not meaningfully enhance that risk. This observed increase in risk associated with α1A-selective ABs compared with 5-ARIs is consistent with biological plausibility. The α1A-AR is expressed in cardiac tissue, and preclinical studies have demonstrated that activation of these receptors protects against multiple forms of injury.^8–19^ Pharmacologic blockade could therefore interfere with these cardioprotective mechanisms. Our findings also align with the ALLHAT trial, which found a 2-fold increased risk of HF with doxazosin compared with chlorthalidone,^20–22^ although that study utilized ABs for hypertension rather than BPH.

### Comparison With Prior Literature

Our cardiovascular event findings are consistent with some prior observational studies. Zhang et al, using Medicare claims data, found that ABs compared with 5-ARIs were associated with increased risk of MACE (HR, 1.08 [95% CI, 1.02-1.13]), though only included one year of follow-up.^68^ Here we have extended follow-up to five years, consistent that these medications typically are taken long term. The prior Zhang et al study used a stricter definition requiring 2 prescriptions within 30 days, whereas our study allowed up to 90 days between prescriptions, which may have included patients with different adherence patterns and hence contributed to the difference in our findings.

Lusty et al found increased risk of HF diagnosis with ABs compared with no BPH medication use, but did not examine other cardiovascular outcomes, did not conduct propensity score-matching and did not compare α1A-selective with non-selective ABs.^69^ A meta-analysis by Sousa et al found ABs associated with increased acute HF risk (OR, 1.78) but did not examine long term risk or other cardiovascular outcomes.^28^ In contrast, the Jackevicius et al VA study found that ABs were not harmful in patients with established HF. This discrepancy may relate to differences in baseline cardiovascular risk between their population (patients with established HF) and ours (patients without prior HF or MI).^29^

### Strengths and Limitations

Our study possesses several strengths. The large sample size from a geographically diverse federated database representing over 158 million patients across 113 healthcare organizations and long term follow-up strengthens the generalizability of our findings.^31–34^ The active comparator, new-user design minimizes confounding by indication and immortal time bias.^39–42^ Propensity score matching achieved excellent balance on numerous cardiovascular risk factors and comorbidities.^41,66^ The consistency of findings across three separate analyses (selective ABs vs 5-ARIs, nonselective ABs vs 5-ARIs, and selective vs nonselective ABs) and across multiple sensitivity analyses strengthens confidence in the observed associations. The use of negative control outcomes supports the specificity of cardiovascular findings.^67^

Several limitations merit consideration. As with all observational studies using electronic health record data, our findings are subject to potential unmeasured confounding and residual confounding by indication.^38,43^ The TriNetX database, while large and diverse, may not be fully representative of all patients with BPH, as it primarily captures patients within academic medical centers and large healthcare systems.^38^ The underrepresentation of Black patients (approximately 6% of our cohort) specifically limits generalizability to this population. Electronic health record–based studies are subject to potential misclassification of exposures and outcomes, although we used validated ICD-10-CM coding algorithms.^35–37,58–64^ The requirement for 3 prescriptions to define medication exposure, while reducing misclassification, may have introduced healthy-adherer bias by selecting patients with better healthcare engagement.

We could not determine the clinical indication for AB prescriptions, as some may have been prescribed for hypertension rather than BPH, though α1A-selective ABs are never prescribed for this indication. Restricting Analysis 1 to selective ABs, which are rarely prescribed for hypertension, partially mitigated this concern. Finally, while we adjusted for numerous potential confounders, the observational nature of this study precludes definitive causal inference.

### Clinical Implications

These findings have potential implications for clinical practice. The consistent association between AB use and increased cardiovascular events, combined with the biological plausibility of α1A-AR blockade interfering with cardioprotective mechanisms,^8–19^ suggests clinicians should consider cardiovascular risk when selecting BPH pharmacotherapy. For patients without established cardiovascular disease, such as those in our study, these findings may inform discussions about BPH treatment options.

ABs remain the preferred first-line therapy for LUTS caused by BPH and they have been shown to improve International Prostate Symptom Score (IPSS) by 5-10 points within 7 days of initiation. Moreover. ABs are well tolerated in most patients, though some can experience orthostatic hypotension, nasal congestion or retrograde ejaculation. In contrast, 5-ARIs can take 3-6 months to elicit symptom relief, with a less robust decline in IPSS than with ABs.^3–7^ Though 5-ARIs offer additional benefits in reducing prostate cancer risk and the need for surgical intervention,^52–54,56^ their use may be further diminished due to their association with significant sexual side effects in up to 8% of men. Given its favorable side effect profile, daily low-dose tadalafil, a phosphodiesterase 5 inhibitor with no recognized cardiovascular risks is being increasingly used as a primary treatment for LUTS.

Shared decision-making that incorporates patient preferences, symptom severity, and consideration of cardiovascular risk profile remains appropriate in the treatment of LUTS due to BPH.

## Conclusions

In this large retrospective cohort study, both α1A-selective and nonselective ABs for BPH were associated with increased risk of cardiovascular events that was evident at 1 year and persisted through 5 years of follow-up. These findings were robust to sensitivity analyses and were specific to cardiovascular endpoints, as negative control outcomes showed no spurious associations. These results may inform shared decision-making for BPH pharmacotherapy. Further investigation, including prospective studies and mechanistic research, is warranted to clarify the cardiovascular effects of ABs in patients with BPH.

## Supplementary Material

**eTable 1.** Baseline demographics for the α1A-AR selective antagonists and non-selective α-blockers groups prior to propensity score matching.

| Category | Characteristic | Cohort Statistics Before PSM |  |  |  |  |  |  |  |
| --- | --- | --- | --- | --- | --- | --- | --- | --- | --- |
| | | Selective α-Blockers | | | 5 - $\alpha$ - Reductase Inhibitors | | | p-value | SMD |
|  |  | Count(%) | Mean | SD | Count(%) | Mean | SD |  |  |
| Demographics | Age at Index | 486285 (100%) | 68.472 | 10.006 | 48098 (100%) | 73.680 | 10.000 | <0.0001 | 0.521 |
|  | Black or African American | 46628 (9.59%) | – | – | 3143 (6.54%) | – | – | <0.0001 | 0.112 |
| Diseases | Essential hypertension | 252487 (51.92%) | – | – | 30016 (62.41%) | – | – | <0.0001 | 0.213 |
|  | Hyperlipidemia | 183643 (37.76%) | – | – | 23691 (49.26%) | – | – | <0.0001 | 0.233 |
|  | Chronic ischemic heart disease | 112413 (23.12%) | – | – | 14912 (31.00%) | – | – | <0.0001 | 0.178 |
|  | Type 2 diabetes | 126467 (26.01%) | – | – | 12439 (25.86%) | – | – | 0.489 | 0.003 |
|  | Chronic kidney disease | 69691 (14.33%) | – | – | 8657 (18.00%) | – | – | <0.0001 | 0.100 |
|  | Sleep apnea | 65884 (13.55%) | – | – | 7146 (14.86%) | – | – | <0.0001 | 0.037 |
|  | Other anemias | 59555 (12.25%) | – | – | 7332 (15.24%) | – | – | <0.0001 | 0.087 |
|  | Cerebrovascular diseases | 61812 (12.71%) | – | – | 7293 (15.16%) | – | – | <0.0001 | 0.071 |
|  | Atherosclerosis | 28904 (5.94%) | – | – | 3240 (6.74%) | – | – | <0.0001 | 0.033 |
|  | Peripheral vascular disease | 29579 (6.08%) | – | – | 3260 (6.78%) | – | – | <0.0001 | 0.028 |
|  | Liver disease | 30416 (6.26%) | – | – | 2780 (5.78%) | – | – | <0.0001 | 0.020 |
|  | Other lung disorders | 16347 (3.36%) | – | – | 1761 (3.66%) | – | – | 0.001 | 0.016 |
|  | HIV disease | 2706 (0.56%) | – | – | 268 (0.56%) | – | – | 0.984 | <0.001 |
| Medications | Beta blockers | 156500 (32.18%) | – | – | 22699 (47.19%) | – | – | <0.0001 | 0.310 |
|  | Aspirin | 129733 (26.68%) | – | – | 18532 (38.53%) | – | – | <0.0001 | 0.255 |
|  | Diuretics | 127928 (26.31%) | – | – | 17587 (36.57%) | – | – | <0.0001 | 0.222 |
|  | Calcium channel blockers | 113894 (23.42%) | – | – | 14833 (30.84%) | – | – | <0.0001 | 0.167 |
|  | ACE inhibitors | 103401 (21.26%) | – | – | 13953 (29.01%) | – | – | <0.0001 | 0.179 |
|  | Angiotensin II inhibitors | 71180 (14.64%) | – | – | 10578 (21.99%) | – | – | <0.0001 | 0.191 |
|  | Vasodilators (cardiac) | 60733 (12.49%) | – | – | 7354 (15.29%) | – | – | <0.0001 | 0.081 |
|  | Hydralazine | 50676 (10.42%) | – | – | 5944 (12.36%) | – | – | <0.0001 | 0.061 |
|  | Carvedilol | 27846 (5.73%) | – | – | 4158 (8.65%) | – | – | <0.0001 | 0.113 |
| Lab Values | Creatinine | 326729 (67.19%) | 1.282 | 2.470 | 37651 (78.28%) | 1.236 | 2.226 | 0.0006 | 0.020 |
|  | Sodium | 320218 (65.85%) | 138.546 | 3.707 | 37268 (77.48%) | 138.819 | 3.450 | <0.0001 | 0.076 |
|  | Potassium | 317338 (65.26%) | 4.203 | 0.494 | 37021 (76.97%) | 4.210 | 0.472 | 0.0097 | 0.014 |
|  | Hemoglobin | 310064 (63.76%) | 13.315 | 2.333 | 35385 (73.57%) | 13.093 | 2.296 | <0.0001 | 0.096 |
|  | Creatinine/Urea ratio | 11438 (2.35%) | 17.481 | 6.456 | 1248 (2.60%) | 18.866 | 6.760 | <0.0001 | 0.210 |
|  | Urea | 2236 (0.46%) | 24.806 | 19.016 | 208 (0.43%) | 28.441 | 17.707 | 0.0081 | 0.198 |

**eTable 2.**
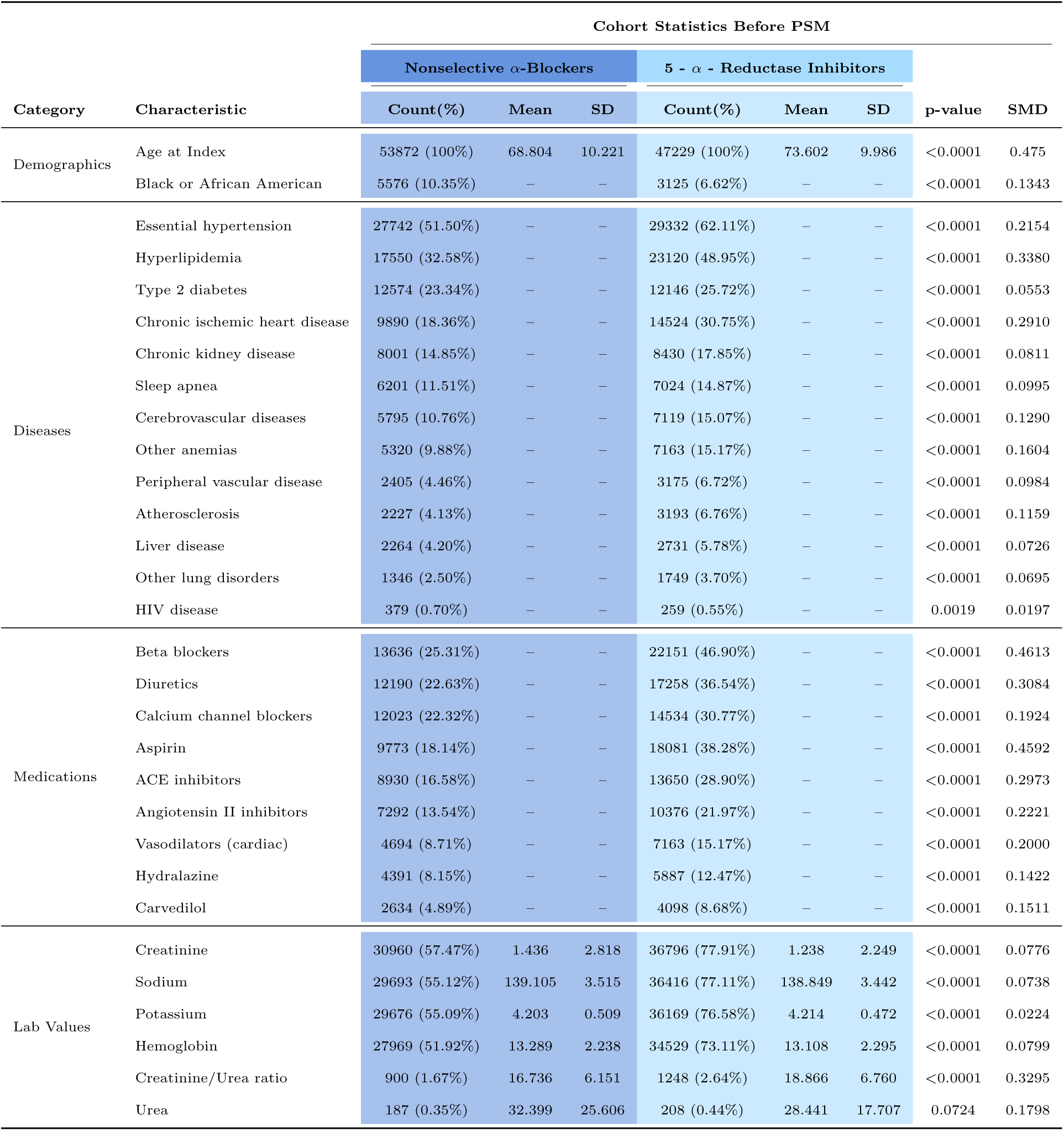
Baseline demographics for the non-selective α-blockers and 5α-reductase inhibitors groups prior to propensity score matching.

|  |  | Cohort Statistics Before PSM |  |  |  |  |  |  |  |
| --- | --- | --- | --- | --- | --- | --- | --- | --- | --- |
| Category | Characteristic | Nonselective α-Blockers | | | 5 - $\alpha$ - Reductase Inhibitors | | | p-value | SMD |
|  |  | Count(%) | Mean | SD | Count(%) | Mean | SD |  |  |
| Demographics | Age at Index | 53872 (100%) | 68.804 | 10.221 | 47229 (100%) | 73.602 | 9.986 | <0.0001 | 0.475 |
|  | Black or African American | 5576 (10.35%) | – | – | 3125 (6.62%) | – | – | <0.0001 | 0.1343 |
| Diseases | Essential hypertension | 27742 (51.50%) | – | – | 29332 (62.11%) | – | – | <0.0001 | 0.2154 |
|  | Hyperlipidemia | 17550 (32.58%) | – | – | 23120 (48.95%) | – | – | <0.0001 | 0.3380 |
|  | Type 2 diabetes | 12574 (23.34%) | – | – | 12146 (25.72%) | – | – | <0.0001 | 0.0553 |
|  | Chronic ischemic heart disease | 9890 (18.36%) | – | – | 14524 (30.75%) | – | – | <0.0001 | 0.2910 |
|  | Chronic kidney disease | 8001 (14.85%) | – | – | 8430 (17.85%) | – | – | <0.0001 | 0.0811 |
|  | Sleep apnea | 6201 (11.51%) | – | – | 7024 (14.87%) | – | – | <0.0001 | 0.0995 |
|  | Cerebrovascular diseases | 5795 (10.76%) | – | – | 7119 (15.07%) | – | – | <0.0001 | 0.1290 |
|  | Other anemias | 5320 (9.88%) | – | – | 7163 (15.17%) | – | – | <0.0001 | 0.1604 |
|  | Peripheral vascular disease | 2405 (4.46%) | – | – | 3175 (6.72%) | – | – | <0.0001 | 0.0984 |
|  | Atherosclerosis | 2227 (4.13%) | – | – | 3193 (6.76%) | – | – | <0.0001 | 0.1159 |
|  | Liver disease | 2264 (4.20%) | – | – | 2731 (5.78%) | – | – | <0.0001 | 0.0726 |
|  | Other lung disorders | 1346 (2.50%) | – | – | 1749 (3.70%) | – | – | <0.0001 | 0.0695 |
|  | HIV disease | 379 (0.70%) | – | – | 259 (0.55%) | – | – | 0.0019 | 0.0197 |
| Medications | Beta blockers | 13636 (25.31%) | – | – | 22151 (46.90%) | – | – | <0.0001 | 0.4613 |
|  | Diuretics | 12190 (22.63%) | – | – | 17258 (36.54%) | – | – | <0.0001 | 0.3084 |
|  | Calcium channel blockers | 12023 (22.32%) | – | – | 14534 (30.77%) | – | – | <0.0001 | 0.1924 |
|  | Aspirin | 9773 (18.14%) | – | – | 18081 (38.28%) | – | – | <0.0001 | 0.4592 |
|  | ACE inhibitors | 8930 (16.58%) | – | – | 13650 (28.90%) | – | – | <0.0001 | 0.2973 |
|  | Angiotensin II inhibitors | 7292 (13.54%) | – | – | 10376 (21.97%) | – | – | <0.0001 | 0.2221 |
|  | Vasodilators (cardiac) | 4694 (8.71%) | – | – | 7163 (15.17%) | – | – | <0.0001 | 0.2000 |
|  | Hydralazine | 4391 (8.15%) | – | – | 5887 (12.47%) | – | – | <0.0001 | 0.1422 |
|  | Carvedilol | 2634 (4.89%) | – | – | 4098 (8.68%) | – | – | <0.0001 | 0.1511 |
| Lab Values | Creatinine | 30960 (57.47%) | 1.436 | 2.818 | 36796 (77.91%) | 1.238 | 2.249 | <0.0001 | 0.0776 |
|  | Sodium | 29693 (55.12%) | 139.105 | 3.515 | 36416 (77.11%) | 138.849 | 3.442 | <0.0001 | 0.0738 |
|  | Potassium | 29676 (55.09%) | 4.203 | 0.509 | 36169 (76.58%) | 4.214 | 0.472 | <0.0001 | 0.0224 |
|  | Hemoglobin | 27969 (51.92%) | 13.289 | 2.238 | 34529 (73.11%) | 13.108 | 2.295 | <0.0001 | 0.0799 |
|  | Creatinine/Urea ratio | 900 (1.67%) | 16.736 | 6.151 | 1248 (2.64%) | 18.866 | 6.760 | <0.0001 | 0.3295 |
|  | Urea | 187 (0.35%) | 32.399 | 25.606 | 208 (0.44%) | 28.441 | 17.707 | 0.0724 | 0.1798 |

**eTable 3.** Baseline demographics for the α1A-AR selective antagonists and non-selective α-blockers groups prior to propensity score matching.

| Category | Characteristic | Cohort Statistics Before PSM |  |  |  |  |  |  |  |
| --- | --- | --- | --- | --- | --- | --- | --- | --- | --- |
| | | α1A-Selective | | | Nonselective α-Blockers | | | p-value | SMD |
|  |  | Count(%) | Mean | SD | Count(%) | Mean | SD |  |  |
| Demographics | Age at Index | 486342 (100%) | 68.471 | 10.005 | 54872 (100%) | 68.847 | 10.233 | <0.0001 | 0.037 |
|  | Black or African American | 46640 (9.59%) | – | – | 5653 (10.30%) | – | – | <0.0001 | 0.024 |
| Diseases | Essential hypertension | 252522 (51.92%) | – | – | 28368 (51.70%) | – | – | 0.319 | 0.004 |
|  | Hyperlipidemia | 183664 (37.76%) | – | – | 17977 (32.76%) | – | – | <0.0001 | 0.105 |
|  | Type 2 diabetes | 126491 (26.01%) | – | – | 12867 (23.45%) | – | – | <0.0001 | 0.059 |
|  | Chronic ischemic heart disease | 112430 (23.12%) | – | – | 10131 (18.46%) | – | – | <0.0001 | 0.115 |
|  | Chronic kidney disease | 69703 (14.33%) | – | – | 8199 (14.94%) | – | – | 0.0001 | 0.017 |
|  | Sleep apnea | 65893 (13.55%) | – | – | 6334 (11.54%) | – | – | <0.0001 | 0.061 |
|  | Cerebrovascular diseases | 61820 (12.71%) | – | – | 5922 (10.79%) | – | – | <0.0001 | 0.060 |
|  | Other anemias | 59566 (12.25%) | – | – | 5440 (9.91%) | – | – | <0.0001 | 0.074 |
|  | Peripheral vascular disease | 29581 (6.08%) | – | – | 2470 (4.50%) | – | – | <0.0001 | 0.071 |
|  | Liver disease | 30424 (6.26%) | – | – | 2301 (4.19%) | – | – | <0.0001 | 0.093 |
|  | Atherosclerosis | 28910 (5.94%) | – | – | 2261 (4.12%) | – | – | <0.0001 | 0.084 |
|  | Other lung disorders | 16350 (3.36%) | – | – | 1353 (2.47%) | – | – | <0.0001 | 0.053 |
|  | HIV disease | 2706 (0.56%) | – | – | 390 (0.71%) | – | – | <0.0001 | 0.019 |
| Medications | Beta blockers | 156515 (32.18%) | – | – | 13975 (25.47%) | – | – | <0.0001 | 0.149 |
|  | Diuretics | 127957 (26.31%) | – | – | 12458 (22.70%) | – | – | <0.0001 | 0.084 |
|  | Calcium channel blockers | 113911 (23.42%) | – | – | 12278 (22.38%) | – | – | <0.0001 | 0.025 |
|  | Aspirin | 129747 (26.68%) | – | – | 9970 (18.17%) | – | – | <0.0001 | 0.205 |
|  | ACE inhibitors | 103410 (21.26%) | – | – | 9142 (16.66%) | – | – | <0.0001 | 0.118 |
|  | Angiotensin II inhibitors | 71189 (14.64%) | – | – | 7429 (13.54%) | – | – | <0.0001 | 0.032 |
|  | Vasodilators (cardiac) | 60736 (12.49%) | – | – | 4797 (8.74%) | – | – | <0.0001 | 0.122 |
|  | Hydralazine | 50687 (10.42%) | – | – | 4464 (8.14%) | – | – | <0.0001 | 0.079 |
|  | Carvedilol | 27851 (5.73%) | – | – | 2676 (4.88%) | – | – | <0.0001 | 0.038 |
| Lab Values | Creatinine | 326779 (67.19%) | 1.282 | 2.470 | 31836 (58.02%) | 1.432 | 2.787 | <0.0001 | 0.057 |
|  | Sodium | 320268 (65.85%) | 138.545 | 3.707 | 30564 (55.70%) | 139.066 | 3.532 | <0.0001 | 0.144 |
|  | Potassium | 317388 (65.26%) | 4.203 | 0.494 | 30541 (55.66%) | 4.200 | 0.509 | 0.198 | 0.008 |
|  | Hemoglobin | 310112 (63.76%) | 13.315 | 2.333 | 28845 (52.57%) | 13.283 | 2.235 | 0.023 | 0.014 |
|  | Creatinine/Urea ratio | 11438 (2.35%) | 17.481 | 6.456 | 900 (1.64%) | 16.736 | 6.151 | 0.0008 | 0.118 |
|  | Urea | 2236 (0.46%) | 24.806 | 19.016 | 187 (0.34%) | 32.399 | 25.606 | <0.0001 | 0.337 |

**eFigure 1.**
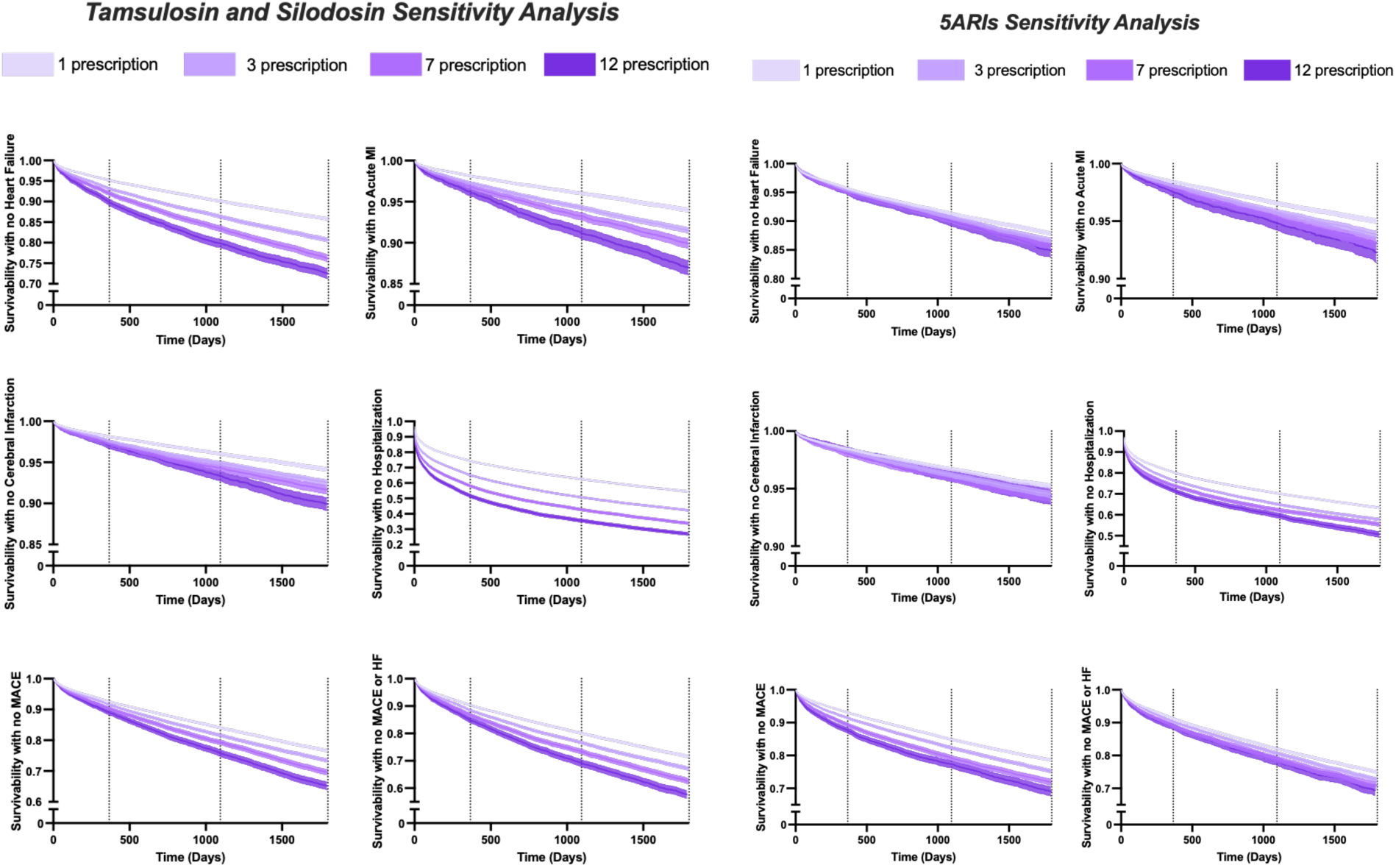
Sensitivity Analysis: Prescription Threshold Variation. Kaplan-Meier curves for all outcomes across varying prescription thresholds (1, 3, 7, and 12 prescriptions) for cohort entry in each analysis.

**eFigure 2.**
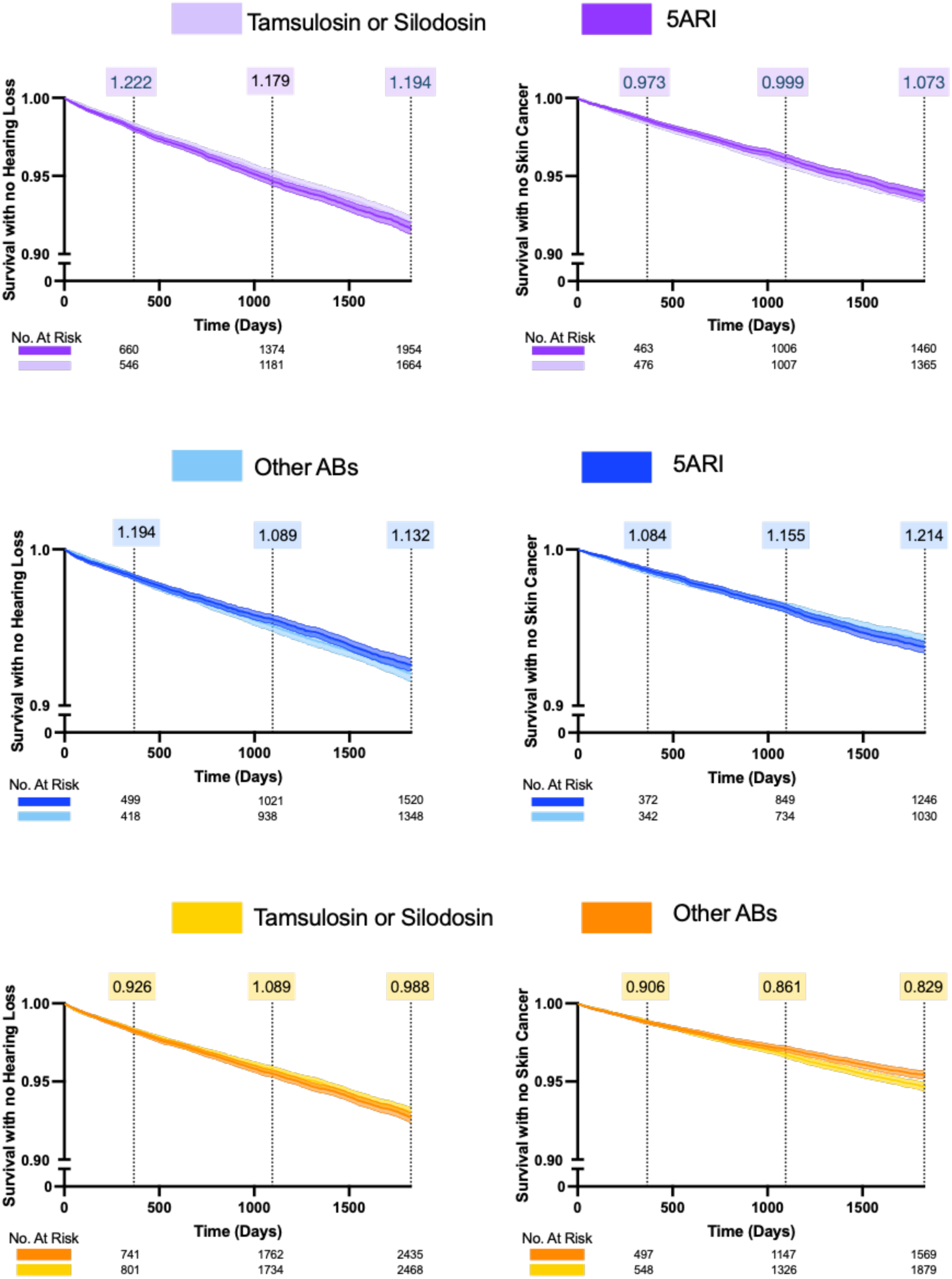
Negative Control Outcomes. Kaplan-Meier curves for sensorineural hearing loss and nonmelanoma skin cancer across all three comparisons.

**eFigure 3.**
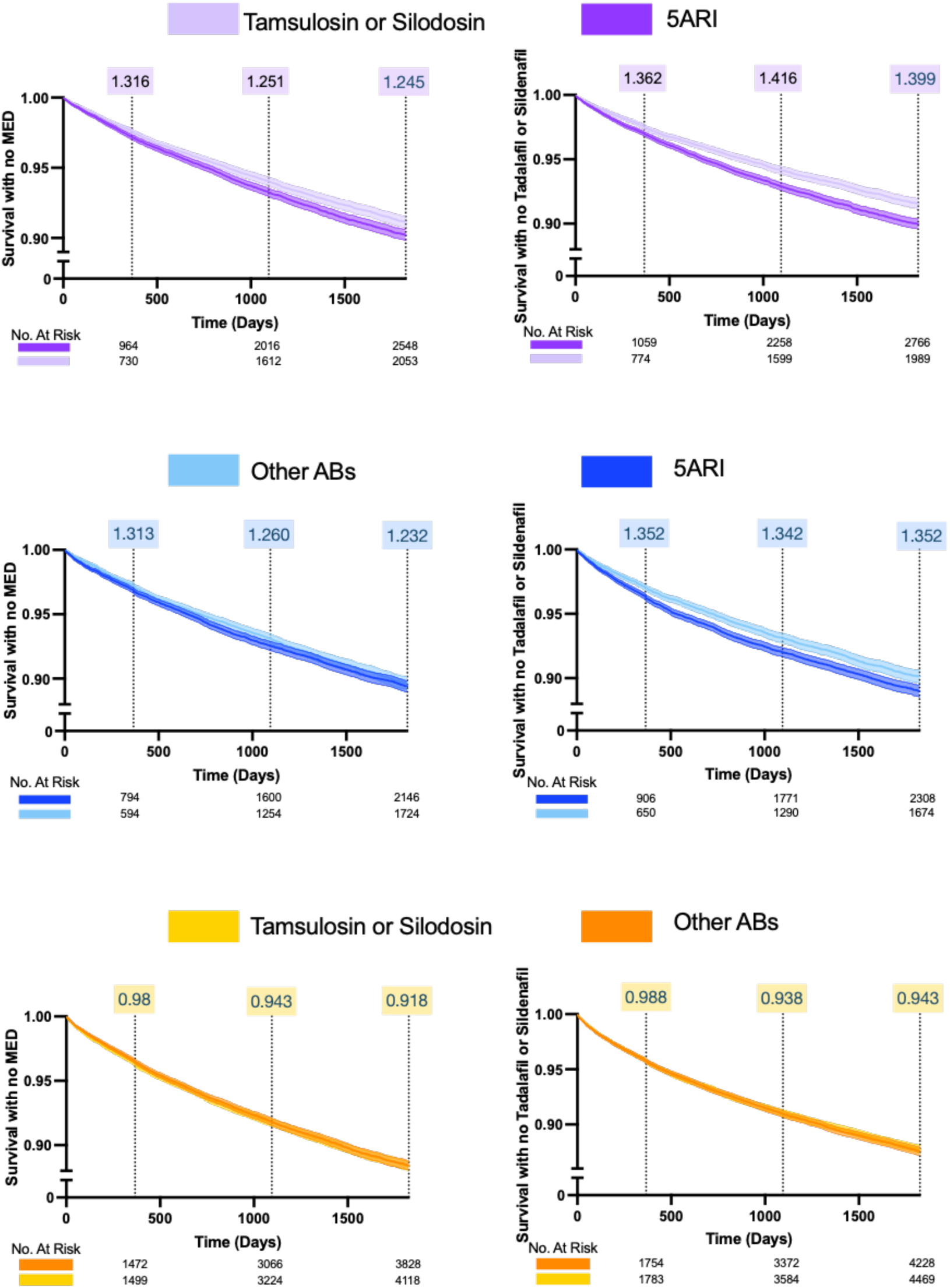
Exploratory Secondary Outcomes. Kaplan-Meier curves for erectile dysfunction and phosphodiesterase-5 inhibitor prescriptions across all three comparisons.

Covariates Included in Analysis

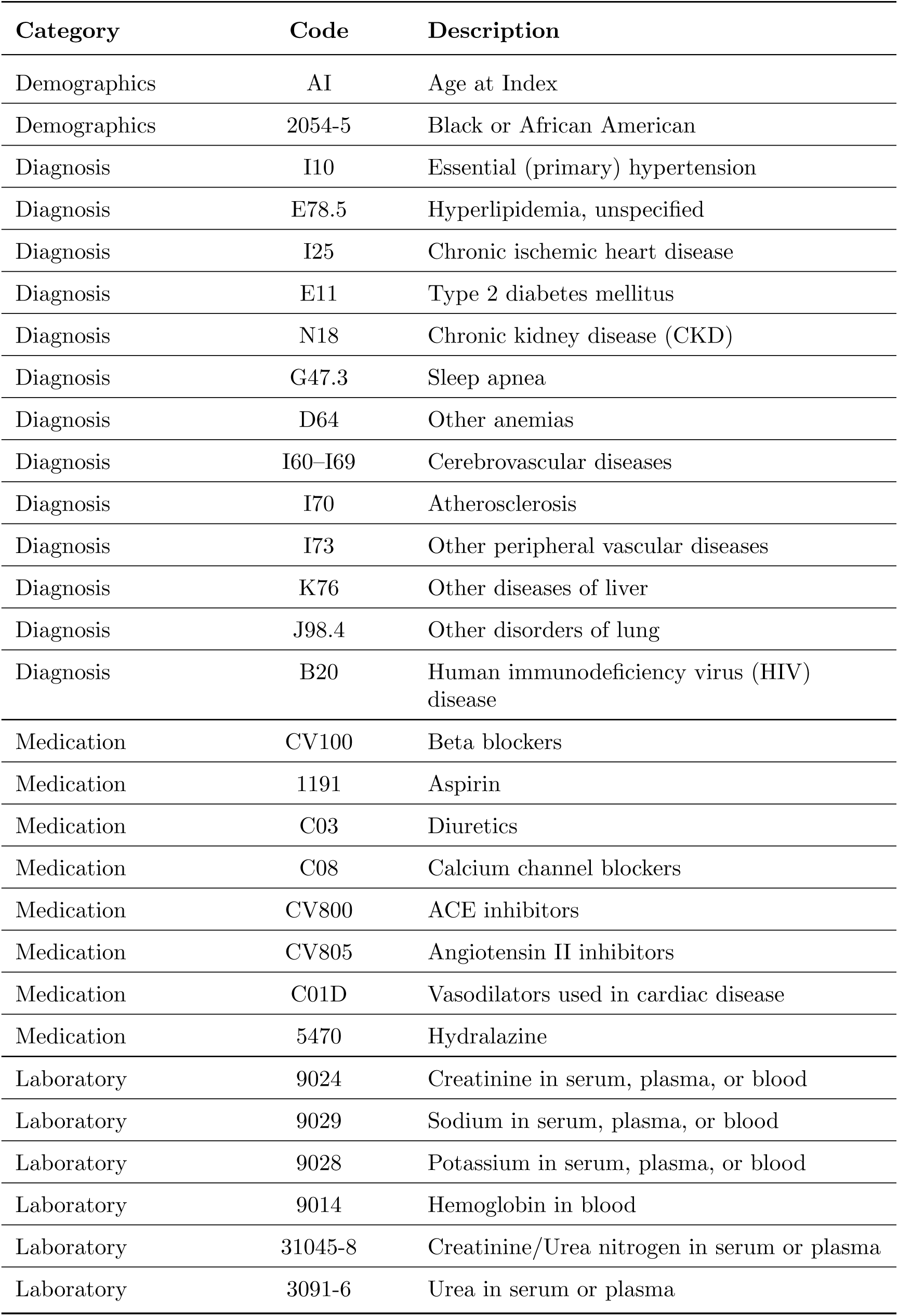

| **Category** | **Code** | **Description** |
| --- | --- | --- |
| Demographics | AI | Age at Index |
| Demographics | 2054-5 | Black or African American |
| Diagnosis | I10 | Essential (primary) hypertension |
| Diagnosis | E78.5 | Hyperlipidemia, unspecified |
| Diagnosis | I25 | Chronic ischemic heart disease |
| Diagnosis | E11 | Type 2 diabetes mellitus |
| Diagnosis | N18 | Chronic kidney disease (CKD) |
| Diagnosis | G47.3 | Sleep apnea |
| Diagnosis | D64 | Other anemias |
| Diagnosis | I60–I69 | Cerebrovascular diseases |
| Diagnosis | I70 | Atherosclerosis |
| Diagnosis | I73 | Other peripheral vascular diseases |
| Diagnosis | K76 | Other diseases of liver |
| Diagnosis | J98.4 | Other disorders of lung |
| Diagnosis | B20 | Human immunodeficiency virus (HIV) disease |
| Medication | CV100 | Beta blockers |
| Medication | 1191 | Aspirin |
| Medication | C03 | Diuretics |
| Medication | C08 | Calcium channel blockers |
| Medication | CV800 | ACE inhibitors |
| Medication | CV805 | Angiotensin II inhibitors |
| Medication | C01D | Vasodilators used in cardiac disease |
| Medication | 5470 | Hydralazine |
| Laboratory | 9024 | Creatinine in serum, plasma, or blood |
| Laboratory | 9029 | Sodium in serum, plasma, or blood |
| Laboratory | 9028 | Potassium in serum, plasma, or blood |
| Laboratory | 9014 | Hemoglobin in blood |
| Laboratory | 31045-8 | Creatinine/Urea nitrogen in serum or plasma |
| Laboratory | 3091-6 | Urea in serum or plasma |

Tamsulosin and Silodosin Group

| # | Criterion | Category | Code | Description |
| --- | --- | --- | --- | --- |
| 1 | At least 18 years old | Demographics | Age | Age (at least 18 years) |
| 2 | Patients with Benign Prostatic Hyperplasia | Diagnosis | N40 | Benign Prostatic Hyperplasia |
| 3 | Not Female (cannot be the following) | Demographics | Gender | Female |
| 4 | Not less than 55 years old (cannot be the following) | Demographics | Age | Age (less than 55 years) |
| 5 | No prostatectomy (cannot have the following) | Procedure | 90470006 | Prostatectomy |
| 6 | No Encounter for Antineoplastic Chemotherapy and Immunotherapy (cannot have the following) | Procedure | Z51.1 | Encounter for Antineoplastic Chemotherapy and Immunotherapy |
| 7 | No Malignant Neoplasm of Prostate (cannot have the following) | Diagnosis | C61 | Malignant Neoplasm of Prostate |
| 8 | No Acquired Absence of Other Genital Organs (cannot have the following) | Diagnosis | Z90.79 | Acquired Absence of Other Genital Organs |
| 9 | No Encounter for Palliative Care (cannot have the following) | Procedure | Z51.5 | Encounter for Palliative Care |
| 10 | Treated with Tamsulosin or Silodosin (one of the following), greater than 3 instances of prescription refills | Medication | 720825 | Silodosin |
|  |  | Medication | 77492 | Tamsulosin |
| 11 | No Testosterone-5-alpha reductase inhibitors treatments (cannot have the following) | Medication | G04CB | Testosterone-5-alpha reductase inhibitors |
| 12 | No other Alpha Blockers (cannot have the following) | Medication | 17300 | Alfuzosin |
|  |  | Medication | 37798 | Terazosin |
|  |  | Medication | 49276 | Doxazosin |
|  |  | Medication | 8629 | Prazosin |

Other Alpha Blockers Group

| # | Criterion | Category | Code | Description |
| --- | --- | --- | --- | --- |
| 1 | At least 18 years old | Demographics | Age | Age (at least 18 years) |
| 2 | Patients with Benign Prostatic Hyperplasia | Diagnosis | N40 | Benign Prostatic Hyperplasia |
| 3 | Not Female (cannot be the following) | Demographics | Gender | Female |
| 4 | Not less than 55 years old (cannot be the following) | Demographics | Age | Age (less than 55 years) |
| 5 | No prostatectomy (cannot have the following) | Procedure | 90470006 | Prostatectomy |
| 6 | No Encounter for Antineoplastic Chemotherapy and Immunotherapy (cannot have the following) | Procedure | Z51.1 | Encounter for Antineoplastic Chemotherapy and Immunotherapy |
| 7 | No Malignant Neoplasm of Prostate (cannot have the following) | Diagnosis | C61 | Malignant Neoplasm of Prostate |
| 8 | No Acquired Absence of Other Genital Organs (cannot have the following) | Diagnosis | Z90.79 | Acquired Absence of Other Genital Organs |
| 9 | No Encounter for Palliative Care (cannot have the following) | Procedure | Z51.5 | Encounter for Palliative Care |
| 10 | Treated with Non-Selective Alpha Blockers (one of the following), greater than 3 instances of prescription refills | Medication | 17300 | Alfuzosin |
|  |  | Medication | 37798 | Terazosin |
|  |  | Medication | 49276 | Doxazosin |
|  |  | Medication | 8629 | Prazosin |
| 11 | No Testosterone-5-alpha reductase inhibitors treatments (cannot have the following) | Medication | G04CB | Testosterone-5-alpha reductase inhibitors |
| 12 | No Tamsulosin or Silodosin (cannot have the following) | Medication | 720825 | Silodosin |
|  |  | Medication | 77492 | Tamsulosin |

5-Alpha Reductase Inhibitors Group

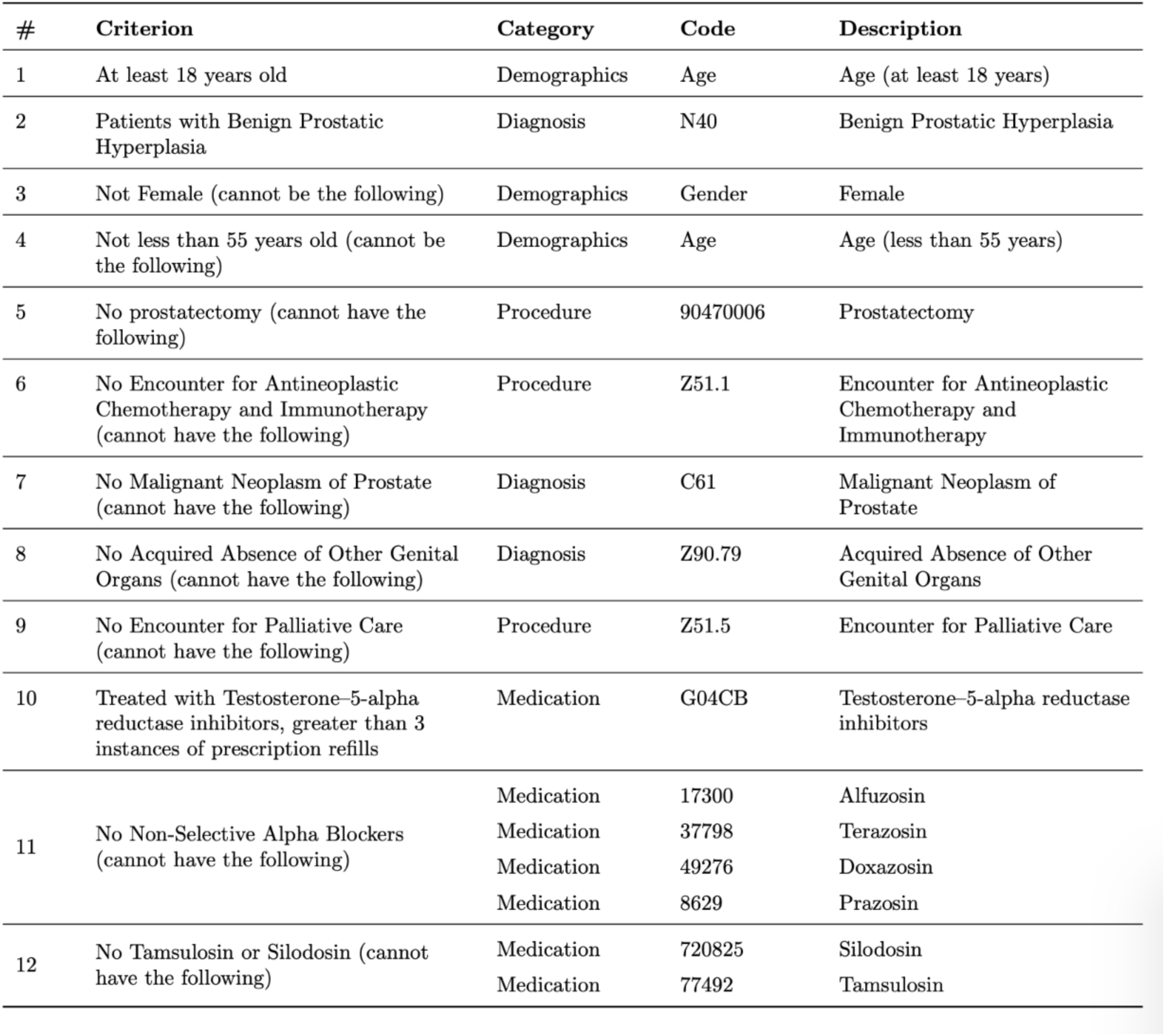

Outcome Definitions

| Outcome | Category | Code | Description |
| --- | --- | --- | --- |
| **Primary Outcomes** |  |  |  |
| Heart Failure | Diagnosis | I50 | Heart Failure |
| Acute Myocardial Infarction | Diagnosis | I21 | Acute Myocardial Infarction |
| Cerebral Infarction | Diagnosis | I63 | Cerebral Infarction |
| Hospitalization | Visit | n/a | Visit: Inpatient Encounter |
|  | Visit | n/a | Visit: Inpatient Non-Acute |
|  | Visit | n/a | Visit: Inpatient Acute |
| MACE (include at least one of the following) | Diagnosis | I21 | Acute Myocardial Infarction |
|  | Diagnosis | I63 | Cerebral Infarction |
|  | Diagnosis | Deceased | Deceased |
| MACE (include at least one of the following) | Diagnosis | I21 | Acute Myocardial Infarction |
|  | Diagnosis | I63 | Cerebral Infarction |
|  | Diagnosis | Deceased | Deceased |
|  | Diagnosis | I50 | Heart Failure |
| **Secondary Outcomes** |  |  |  |
| Male Erectile Dysfunction | Diagnosis | N52 | Male Erectile Dysfunction |
| MACE (include at least one of the following) | Medication | 358263 | Tadalafil |
|  | Medication | 136411 | Sildenafil |
| **Negative Outcomes** |  |  |  |
| Sensorineural Hearing Loss | Diagnosis | H90 | Conductive and sensorineural hearing loss |
| Skin Cancer | Diagnosis | C44 | Other and unspecified malignant neoplasm of skin |

\*\*Patients with the outcome of interest occurring prior to the index event (defined as the third prescription refill) were excluded from outcome analyses.\*\*

## Data Availability

All data will be available upon request after publication.

